# Parental migration, socioeconomic deprivation and hospital admissions in preschool children: national cohort study

**DOI:** 10.1101/2024.01.22.24301591

**Authors:** Kate M Lewis, Rachel Burns, Mario Cortina-Borja, Anja Heilmann, Alison Macfarlane, Selina Nath, Sarah Salway, Sonia Saxena, Nazmy Villarroel-Williams, Russell Viner, Pia Hardelid

## Abstract

**Background:** A third of children born in England have at least one parent born outside the UK, yet family migration history is infrequently studied as a social determinant of child health. We describe differences in rates of hospital admissions in children aged up to five years by parental migration and socioeconomic group.

**Methods:** Birth registrations linked to Hospital Episode Statistics were used to derive a cohort of 4,174,596 children born in state-funded hospitals in England between 2008 and 2014, with follow-up until age five years. We looked at eight maternal regions of birth, maternal country of birth for the 6 most populous groups and parental migration status for the mother and second parent (UK-born/non-UK-born). We used Index of Multiple Deprivation (IMD) quintiles to indicate socioeconomic deprivation. We fitted negative binomial/Poisson regression models to model associations between parental migration groups and the risk of hospital admissions, including interactions with IMD group.

**Findings:** Children of UK-born (73.6% of the cohort) mothers had the highest rates of emergency admissions (171.6 per 1000 child-years, 95% confidence interval (CI) 171.4-171.9), followed by South Asia-born mothers (155.9 per 1000, 95% CI 155.1-156.7). The high rates estimated in the South Asia group were driven by children of women born in Pakistan (186.8 per 1000, 95% CI 185.4, 188.2). A socioeconomic gradient in emergency admissions was present across all maternal region of birth groups, but most pronounced among children of UK-born mothers (incidence rate ratio 1.43, 95% CI 1.42-1.44, high vs. low IMD group). Overall, children whose parents were both born abroad had lower emergency admission rates than children whose parents were both born in the UK. Patterns of planned admissions followed a similar socioeconomic gradient and were highest among children with mothers born in Middle East and North Africa, and South Asia.

**Interpretation:** This research indicates that children whose parents who have migrated to the UK generally have lower overall usage of NHS emergency inpatient services than children of UK-born parents. Our study revealed a socioeconomically graded patterns of hospital admissions for all children born in England, which were highest amongst those with mothers born in the UK, South Asia, and the Middle East and North Africa. Future research using linked primary and secondary care datasets will improve understanding on whether healthcare use is proportionate to need.

**Funding:** National Institute for Health Research.

## Introduction

International migration, defined here as the movement of people to countries outside their place of birth, is a growing feature of our modern globalised world.^1^ The reasons for international migration are multifaceted, including push factors such as conflict, political instability and the climate crisis, and pull factors, such as employment, study opportunities, and network/family ties.^1–3^ Likewise, the health of international migrants and their children are also highly heterogeneous, shaped not only by structural and political factors in their destination country, but by those that led to departure from their home country and encountered in transit countries.^1^ Nonetheless, people who migrate face shared challenges navigating new health and social care services for themselves and their children, which, for some migrant groups in particular, may operate within legislation hostile to their presence.^4^ In conjunction with other interlinking social determinants of health, parental migration is, therefore, an important topic of public health inquiry.^1,5–7^

A third (34.2%) of babies born in England and Wales in 2021 had at least one parent born outside the UK.^8^ Despite the size of this population, and the increased propensity for these children to be living in poverty,^9^ there is no national-level research on the health and healthcare utilisation of children born to migrant parents in the UK.^2^ This reflects the predominant focus on ethnicity, rather than migration history or country of birth, as a determinant of health in the UK and, relatedly, the lack of recording of migration status in routine health data.^7^ A systematic review on health service use, mostly conducted in North America and Europe (but no UK-based studies), identified higher hospital and emergency service use among first and second generation migrant children compared with the rest of the childhood population.^6^ This suggests that migrants and their children face barriers in accessing appropriate and timely healthcare. However, differences in migrant populations, health systems, and health outcomes, limit the generalisability of these findings to the UK.

The aim of this study is to describe the combined association of parental migration (defined as maternal world region/country of birth and parental migration status) and socioeconomic deprivation with rates of early childhood hospital admissions. Our particular focus will be on maternal region of birth. We compare rates of emergency and planned hospital admissions overall and for three common childhood conditions (acute infections, feeding difficulties and jaundice, and tooth extractions for caries), which are considered preventable with the appropriate care provision.

## Methods

### Data sources and linkage

To conduct this population-based cohort study, we used linked Office for National Statistics (ONS) birth and ONS death registration data, and Hospital Episode Statistics Admitted Patient Care (HES APC) records from England (Table 1). Birth and death registrations are routinely linked by ONS using a deterministic matching algorithm. This study uses three types of HES APC records: maternal deliveries, infant births and re-admissions. Linkage between the ONS and HES datasets was carried out by NHS Digital in partnership with the ONS and City, University of London as part of a previous NIHR funded study.^10^ The deterministic stepwise algorithm used to link these datasets, and quality assurance methods, are described elsewhere.^10–12^

**Table 1.**
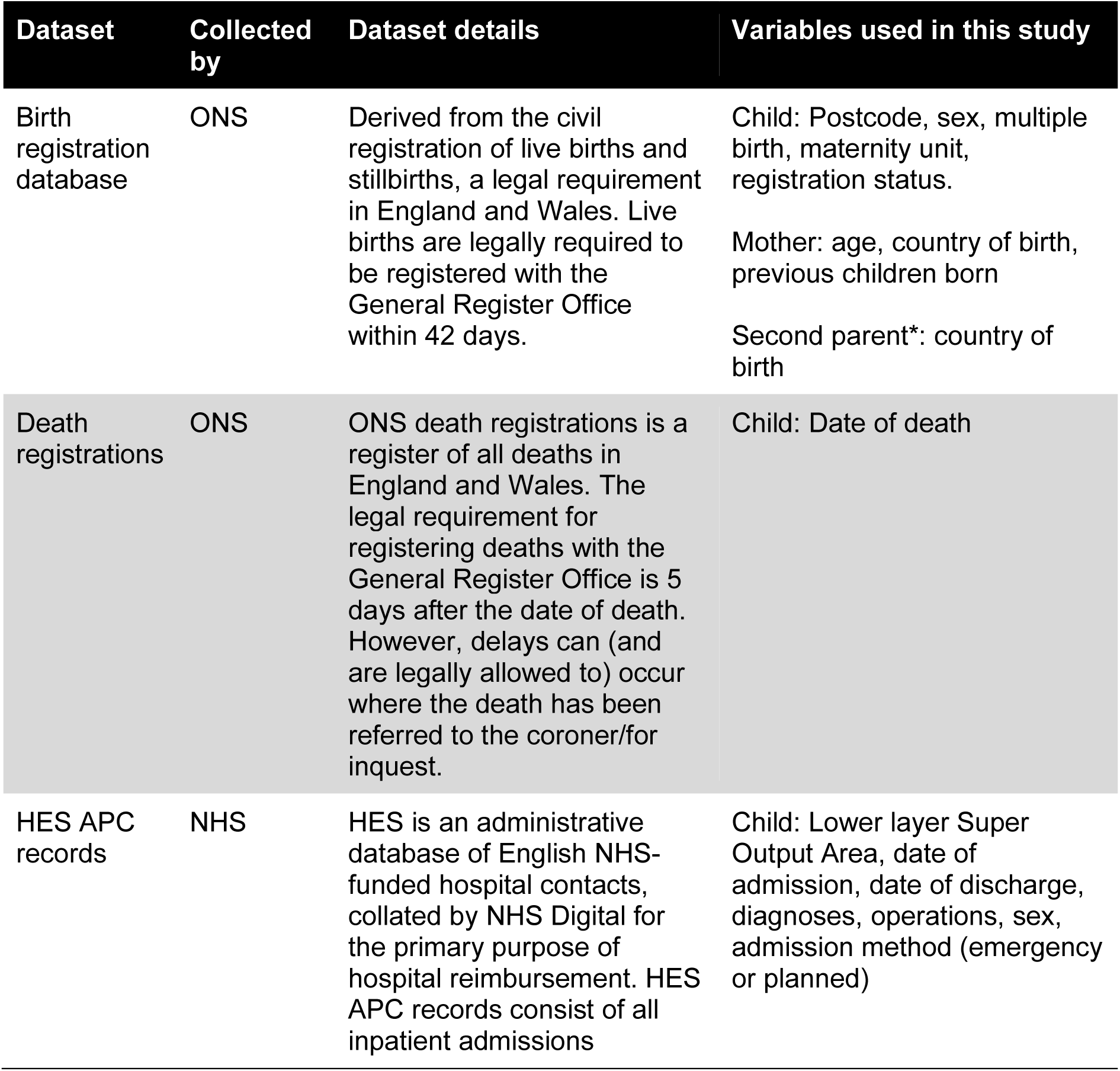
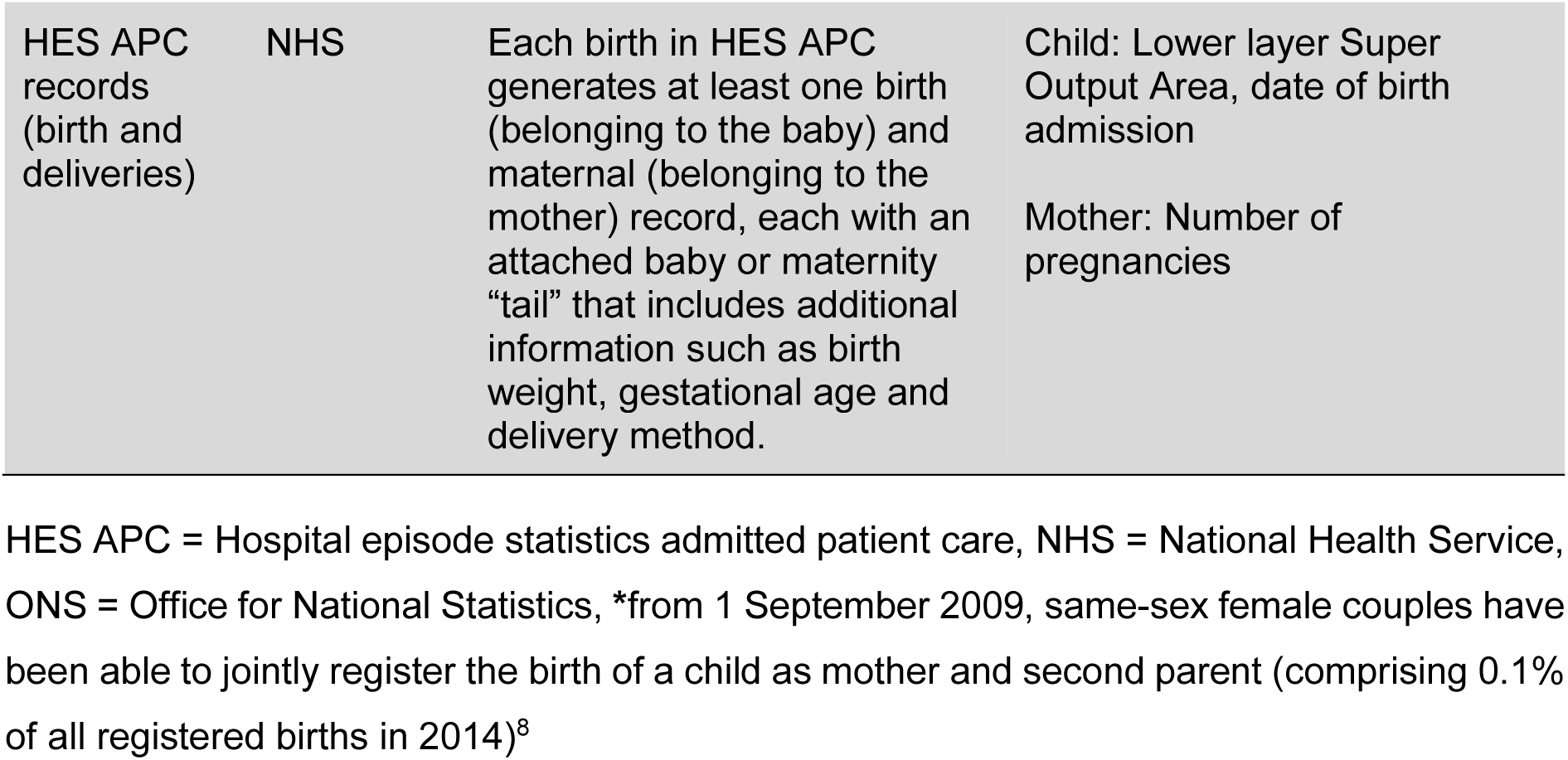
Data sources and variables used in the population based cohort study.

| **Dataset** | **Collected by** | **Dataset details** | **Variables used in this study** |
| --- | --- | --- | --- |
| Birth registration database | ONS | Derived from the civil registration of live births and stillbirths, a legal requirement in England and Wales. Live births are legally required to be registered with the General Register Office within 42 days. | Child: Postcode, sex, multiple birth, maternity unit, registration status.<br>Mother: age, country of birth, previous children born<br>Second parent*: country of birth |
| Death registrations | ONS | ONS death registrations is a register of all deaths in England and Wales. The legal requirement for registering deaths with the General Register Office is 5 days after the date of death. However, delays can (and are legally allowed to) occur where the death has been referred to the coroner/for inquest. | Child: Date of death |
| HES APC records | NHS | HES is an administrative database of English NHS-funded hospital contacts, collated by NHS Digital for the primary purpose of hospital reimbursement. HES APC records consist of all inpatient admissions | Child: Lower layer Super Output Area, date of admission, date of discharge, diagnoses, operations, sex, admission method (emergency or planned) |
| HES APC records (birth and deliveries) | NHS | Each birth in HES APC generates at least one birth (belonging to the baby) and maternal (belonging to the mother) record, each with an attached baby or maternity “tail” that includes additional information such as birth weight, gestational age and delivery method. | Child: Lower layer Super Output Area, date of birth admission<br>Mother: Number of pregnancies |

HES APC = Hospital episode statistics admitted patient care, NHS = National Health Service, ONS = Office for National Statistics, \*from 1 September 2009, same-sex female couples have been able to jointly register the birth of a child as mother and second parent (comprising 0.1% of all registered births in 2014)<sup>8</sup>

### Study cohort

We included live births between 1^st^ January 2008 and 31^st^ December 2014 in England. Infants were excluded if they were: from a multiple birth (identified using birth registration records) because of an increased risk of false matches in HES records;^13^ born to mothers not resident in England (identified using resident postcode at delivery on birth registration records) to prevent systematic loss to follow up; or born in a military or private maternity unit (identified using the maternity unit identifier on birth registration records) as these children did not have accompanying HES APC birth record. ONS birth registrations unlinked to a HES APC birth record were also excluded in our study, as the cohort had been configured such that babies without a HES APC birth record did not have a link to HES APC admission data, therefore these children cannot be followed up. Characteristics of children with and without linked records are explored in the results section. Home births were included in this study if they had linked birth registration-HES APC records. Finally, children with missing place of residence, missing maternal place of birth, or with ≤1 day follow-up were excluded. Follow-up start and end date differ by outcome, as shown in Table 2. The maximum available follow-up date was 31^st^ December 2014.

**Table 2.**
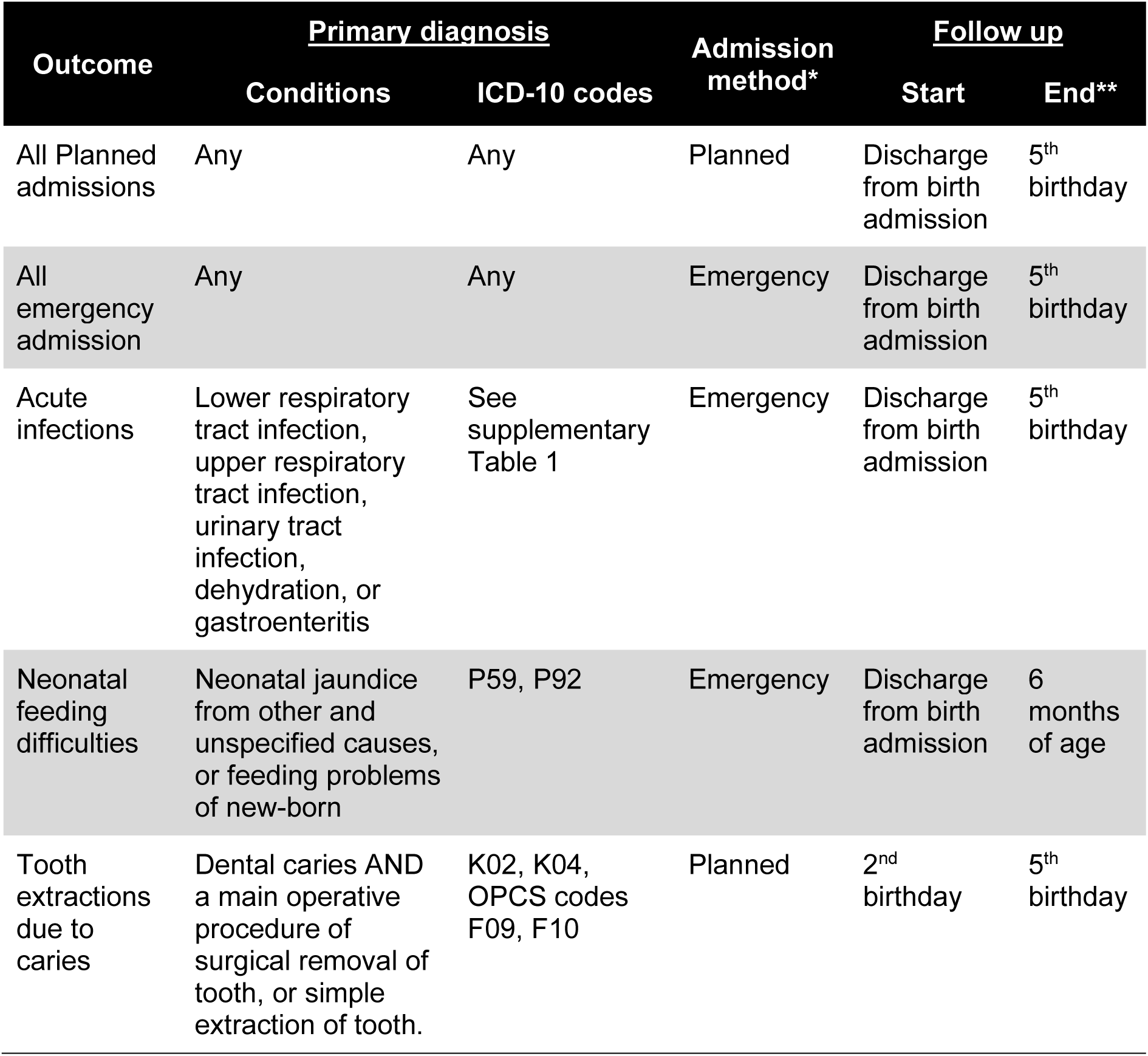
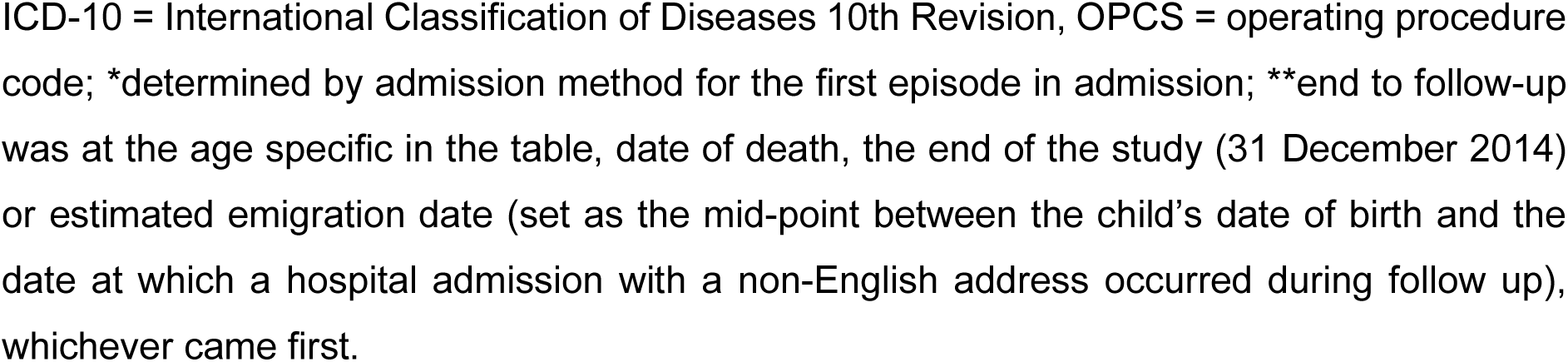
Case definitions and follow up time for each study outcome.

| Outcome | <u>Primary diagnosis</u> |  | Admission method* | <u>Follow up</u> |  |
| --- | --- | --- | --- | --- | --- |
|  | Conditions | ICD-10 codes |  | Start | End** |
| All Planned admissions | Any | Any | Planned | Discharge from birth admission | 5<sup>th</sup> birthday |
| All emergency admission | Any | Any | Emergency | Discharge from birth admission | 5<sup>th</sup> birthday |
| Acute infections | Lower respiratory tract infection, upper respiratory tract infection, urinary tract infection, dehydration, or gastroenteritis | See supplementary Table 1 | Emergency | Discharge from birth admission | 5<sup>th</sup> birthday |
| Neonatal feeding difficulties | Neonatal jaundice from other and unspecified causes, or feeding problems of new-born | P59, P92 | Emergency | Discharge from birth admission | 6 months of age |
| Tooth extractions due to caries | Dental caries AND a main operative procedure of surgical removal of tooth, or simple extraction of tooth. | K02, K04, OPCS codes F09, F10 | Planned | 2<sup>nd</sup> birthday | 5<sup>th</sup> birthday |

ICD-10 = International Classification of Diseases 10th Revision, OPCS = operating procedure code; \*determined by admission method for the first episode in admission; \*\*end to follow-up was at the age specific in the table, date of death, the end of the study (31 December 2014) or estimated emigration date (set as the mid-point between the child's date of birth and the date at which a hospital admission with a non-English address occurred during follow up), whichever came first.

### Outcomes

There were five hospital admission-based outcomes in this study (Table 2). We defined an admission as one continuous stay at a hospital, including admissions within one day of each other and transfers between hospitals.^13^ We first examined incidence rates of admissions up to age five years irrespective of diagnosis, stratified by admission method (emergency or planned). Emergency admissions are defined by NHS England as “ unpredictable and at short notice because of clinical need” and planned (or elective) admissions defined as occurring where “the decision to admit could be separated in time from the actual admission”.^14^ Our secondary outcomes included three common causes of paediatric hospital admissions that may be avoidable with preventative or responsive primary/community care:^15–17^ emergency admissions for acute infections up to age five years, emergency admissions for feeding problems or jaundice up to six months; and planned admissions for tooth extraction due to caries from two to five years. Reducing childhood admissions for lower respiratory tract infections and tooth extractions for caries have been specifically identified by the Department of Health and Social Care as priorities within the NHS Outcomes Framework.^2^ Hospital admissions due to neonatal feeding problems may be preventable with feeding advice and support, in the immediate post-neonatal period, either in the hospital after delivery, or at home.^3^

### Exposures

The primary exposure in this study was maternal world region of birth, derived from mother’s country of birth recorded at birth registration. This was categorised into eight groups, adapted from the World Bank’s 7-group classification of geographical regions (separating the UK from the Europe and Central Asia group, see Figure 1).^18^ We also present results by maternal countries of birth for the six most common countries in our dataset (Bangladesh, India, Nigeria, Pakistan, Poland and the UK). A six-category parental “migration status” variable was also created as a secondary exposure, defined by mothers’ and second parents’ countries of birth grouped into UK, non-UK and sole mother registration, where second parent information was not recorded.

**Figure 1.**
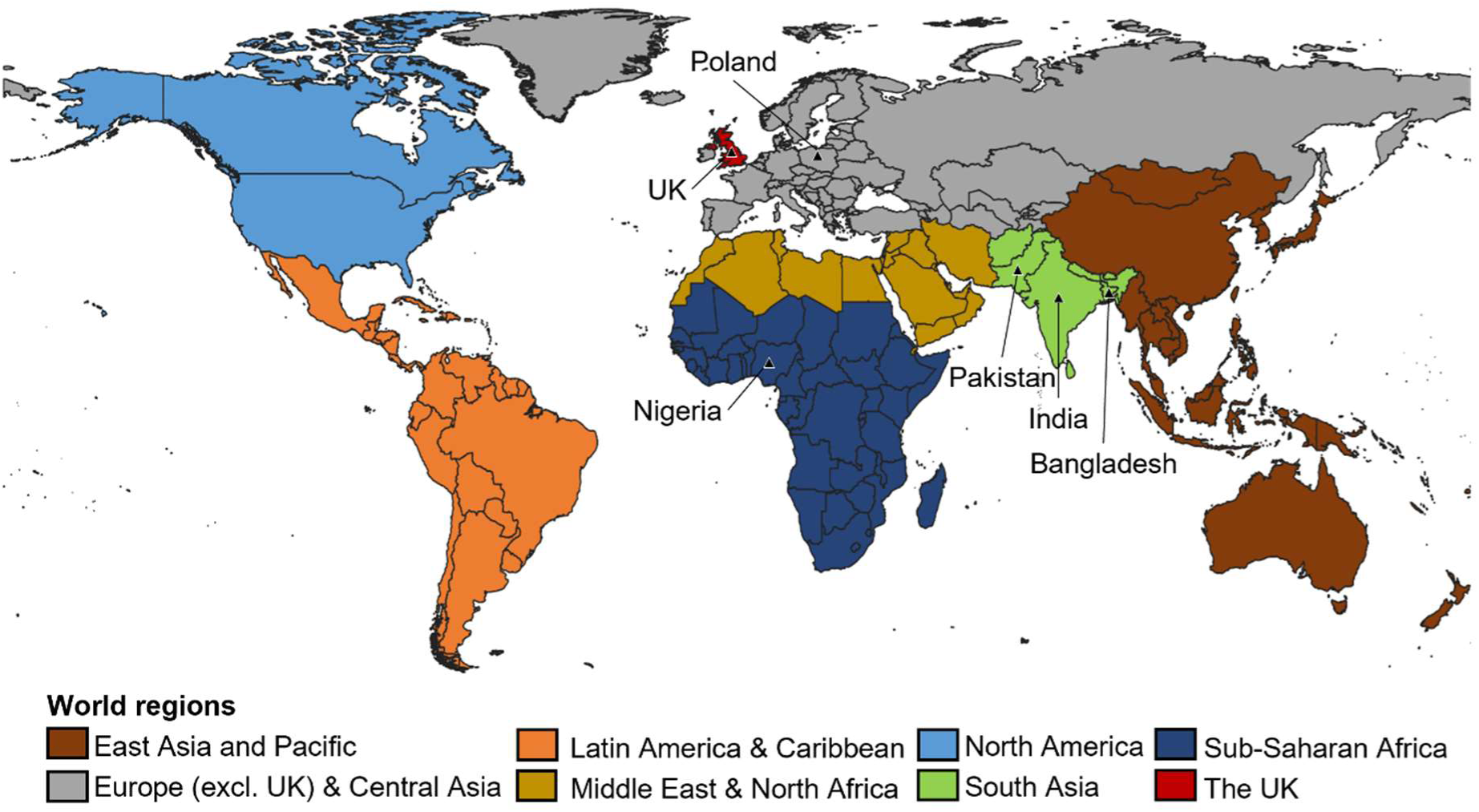
Categories of maternal world regions of birth (with countries of birth indicated by the triangular symbols)

Socioeconomic deprivation was measured using the Index of Multiple Deprivation (IMD) 2010.^19^ IMD is constructed using seven domains of deprivation (income, employment, education and skills, health and disability, crime, barriers to housing and services, and living environment) calculated at the Lower layer Super Output Area level, an area with an average population of 1500 residents or 650 households. Scores are weighted and ranked to produce a relative measure of deprivation for each small area across England. In this study, IMD is based on each child’s residential postcode at delivery from birth registration data (supplemented by HES APC birth or delivery records if missing), and is split into 5 groups (from most to least deprived). Whilst IMD is a small-area level measure of deprivation, we used it as an indicator of family-level socioeconomic position in this study (a common approach to health disparities research in the UK).^20^

### Covariates

We also selected the following variables to be summarised given their association with migration and/or hospital admission rates as shown in the research literature: maternal self-reported ethnicity, child’s sex, region of residence, presence of congenital anomaly, year of birth and maternal age (see Supplementary Table 2 for definitions).^1,17,21,22^ To help guide the selection of confounders for our final statistical models, we constructed a directed acyclic graph (DAG) of variables that are related to parental migration and child hospital admissions. Supplementary Figure 1 shows a simplified DAG of maternal country of birth and child emergency admissions. We included only year of birth in the statistical models as we hypothesise that the other covariates occur temporally after the exposure and therefore cannot be confounders of the relationship between parental migration and hospital admission. We did not include maternal ethnicity in the models due to high levels of missingness in this variable and the potential of introducing overadjustment bias into the results (owing to the overlaps between facets encompassing ethnicity and country of birth e.g. culture and traditions, language, religion).^23^

### Statistical methods

We described the distribution of key childhood characteristics by maternal world region of birth. Unadjusted incidence rates of hospital admissions for children within the cohort were then calculated by dividing the number of admissions by person-time at risk per 1000 child-years.

To examine whether incidence rates of admissions varied by maternal world region of birth and IMD group, we fitted separate regression models for each outcome with the logarithm of person-time as the offset. We used negative binomial or Poisson regression models depending on the presence of overdispersion in each model as indicated by Pearson’s dispersion statistic. To account for unmeasured shared factors of children in the cohort with the same mother, we used robust standard errors clustered at the family level. Alongside maternal world region of birth and IMD group, we adjusted for year of birth to account for changes over time, including population composition, thresholds for hospital admission and infection risk.^24^ We included interaction terms between maternal world region of birth and IMD groups to model the combined association of both exposures, comparing models’ specifications with the Akaike information criterion value (AIC; with a smaller value indicating preferable model fit). We estimated the adjusted incidence rate ratios (IRRs) of admission for each IMD group in comparison to the least deprived IMD group, within maternal region groups. We further estimated marginal incidence rates of hospital admissions for each level of the interaction between maternal region of birth and IMD group, with year of birth set to mid-study (2011). We repeated the above analysis by maternal country of birth and parental migration status.

Two sensitivity analyses were performed. We repeated the above analyses stratifying the cohort by London or non-London region of birth to account for known geographical variability in migrant populations and incidence rates of hospital admissions, particularly in differential thresholds for admissions.^21^ Secondly, since data on embarkations from the UK were not available and very few emigrations are captured in HES, we ran simulations to assess how different levels of international emigration could affect the final results (see Appendix C for details).

Statistical analyses were conducted in Stata v16 within the ONS Secure Research Service.

### Patient and public involvement

A group of women (*n*=7), born in a range of countries outside the UK, who had children whilst living in the UK, took part in a group discussion about our research at the beginning stages of this project. The group provided feedback on our project objectives and avenues for future research directions, as well sharing personal insights from their own experiences of navigating the NHS healthcare system in pregnancy and the early years.

## Results

There were 4,736,499 live births in England between 1 January 2008 and 31 December 2014 recorded in the ONS birth registration dataset. As shown in Figure 2, 175,834 (3.7%) of these births met at least one of the predefined exclusion criteria and an additional 370,730 (8.1%) birth registrations were also excluded as they did not have an associated HES APC birth record. Patterns of the missing 8.1% HES APC birth records differed across child characteristics, and was particularly common in: earlier study years (14.9% in 2008 vs 6.2% in 2014); children with mothers born in North America (9.6%), Latin America and Caribbean (9.2%) and Sub-Saharan Africa (9.1%); children without joint birth registrations (12.6% for UK-born mothers and 9.9% for non-UK-born-mothers); and the North West of England (9.7% compared to 3.8% in North East) (see Supplementary Table 5). Lastly, 15,336 (0.4%) linked records were excluded because of missing information on residential address, mothers place of birth, or ≤1 day of follow up time.

**Figure 2.**
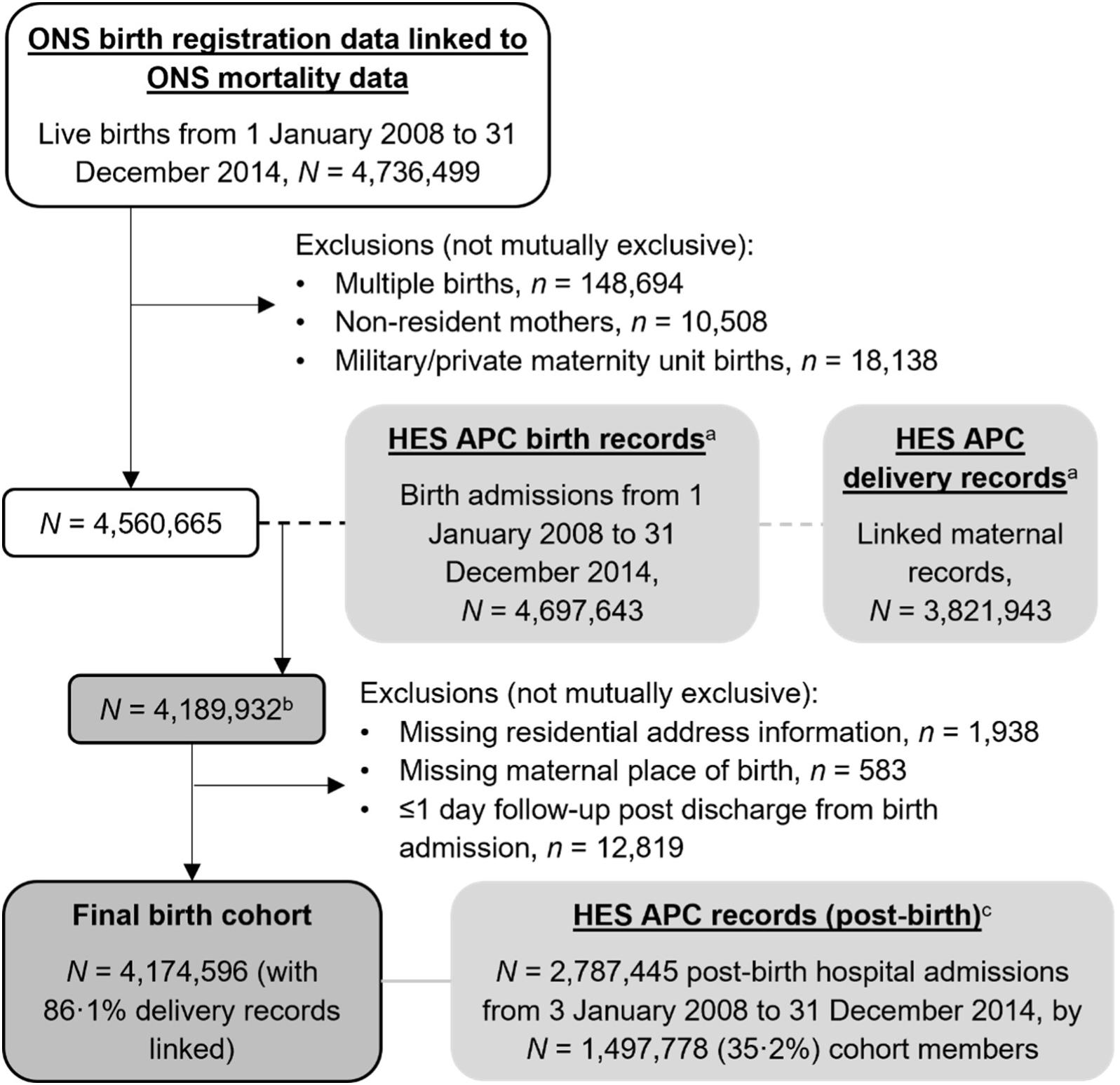
Flow chart showing study sample derivation; HES APC = Hospital Episode Statistics Admitted Patient Care, ONS = Office for National Statistics ^a^Data opt outs applied (<u>digital.nhs.uk/services/national-data-opt-out</u>); ^b^Linkage with 91.9% of ONS birth registrations after exclusions; ^c^Any admission to hospital after discharge from birth admission to 5 years of age within study period

### Cohort characteristics

The final cohort included 4,174,596 children (48.7% female), of whom 1,100,827 (26.4%) had mothers born outside the UK. The most common maternal countries of birth, after the UK, were Poland (122,889; 2.9% of the cohort), Pakistan (115,886; 2.8%), India (85,830; 2.1%), Bangladesh (50,831; 1.2%), and Nigeria (42,934, 1.0%; Supplementary Table 6). A non-UK-born second parent was recorded for 1,025,992 (24.6%) children and 233,864 (5.6%) had no second parent recorded on their birth registration (6.1% with UK-born mothers compared to 4.1% with non-UK-born mothers, Table 3). In total, 1,292,853 children (31.0% of those with records for two parents) had at least one parent born outside the UK. More than 40% of children with mothers born in Sub-Saharan Africa, the Middle East and North Africa, and South Asia were in the most deprived IMD group, compared to 24.5% whose mothers were born in the UK. 11.0% of children with mothers born in the UK had a London-based residence at birth, compared with 36.3%-59.9% of mother born outside the UK. Mothers born in East Asia and Pacific, and North America tended to be older at delivery and the proportion of children identified as having a congenital anomaly was highest in those with mothers born in South Asia, followed by the UK (2.8% and 2.7%, respectively).

**Table 3.** Key characteristics of final study cohort, by maternal world region of birth.

|  | East-Asia &<br>Pacific<br>N (%) | Europe &<br>Central Asia<br>N (%) | Latin<br>America &<br>Caribbean<br>N (%) | Middle East &<br>North Africa<br>N (%) | North America<br>N (%) | South Asia<br>N (%) | Sub-Saharan<br>Africa<br>N (%) | UK<br>N (%) | Total<br>N (%) |
| --- | --- | --- | --- | --- | --- | --- | --- | --- | --- |
| Total | 94158 (100) | 380504 (100) | 40413 (100) | 60820 (100) | 22534 (100) | 296976 (100) | 205422 (100) | 3073769 (100) | 4174596 (100) |
| SP place of birth |  |  |  |  |  |  |  |  |  |
| UK | 37227 (39.5) | 98791 (26.0) | 12351 (30.6) | 7656 (12.6) | 14511 (64.4) | 64931 (21.9) | 31394 (15.3) | 2647879 (86.1) | 2914740 (69.8) |
| Non-UK | 54746 (58.1) | 264876 (69.6) | 24624 (60.9) | 52110 (85.7) | 7663 (34.0) | 229779 (77.4) | 154718 (75.3) | 237476 (7.7) | 1025992 (24.6) |
| No SP registered | 2185 (2.3) | 16837 (4.4) | 3438 (8.5) | 1054 (1.7) | 360 (1.6) | 2266 (0.8) | 19310 (9.4) | 188414 (6.1) | 233864 (5.6) |
| Year of birth |  |  |  |  |  |  |  |  |  |
| 2008 | 12992 (13.8) | 42017 (11.0) | 5532 (13.7) | 7527 (12.4) | 2942 (13.1) | 39587 (13.3) | 29338 (14.3) | 410573 (13.4) | 550508 (13.2) |
| 2009 | 13462 (14.3) | 47054 (12.4) | 5903 (14.6) | 8545 (14.0) | 3081 (13.7) | 41158 (13.9) | 30113 (14.7) | 434200 (14.1) | 583516 (14.0) |
| 2010 | 13987 (14.9) | 52493 (13.8) | 5859 (14.5) | 8835 (14.5) | 3283 (14.6) | 41600 (14.0) | 30573 (14.9) | 452316 (14.7) | 608946 (14.6) |
| 2011 | 13631 (14.5) | 54812 (14.4) | 5711 (14.1) | 8495 (14.0) | 3254 (14.4) | 43580 (14.7) | 29737 (14.5) | 455068 (14.8) | 614288 (14.7) |
| 2012 | 14491 (15.4) | 59260 (15.6) | 5854 (14.5) | 8986 (14.8) | 3353 (14.9) | 45172 (15.2) | 30043 (14.6) | 457820 (14.9) | 624979 (15.0) |
| 2013 | 12825 (13.6) | 61065 (16.0) | 5751 (14.2) | 9149 (15.0) | 3262 (14.5) | 43879 (14.8) | 28510 (13.9) | 436532 (14.2) | 600973 (14.4) |
| 2014 | 12770 (13.6) | 63803 (16.8) | 5803 (14.4) | 9283 (15.3) | 3359 (14.9) | 42000 (14.1) | 27108 (13.2) | 427260 (13.9) | 591386 (14.2) |
| IMD group |  |  |  |  |  |  |  |  |  |
| 1 Least deprived | 15269 (16.2) | 42101 (11.1) | 3859 (9.5) | 4142 (6.8) | 5288 (23.5) | 15405 (5.2) | 15866 (7.7) | 527475 (17.2) | 629405 (15.1) |
| 2 | 15904 (16.9) | 52849 (13.9) | 4393 (10.9) | 5922 (9.7) | 5385 (23.9) | 22930 (7.7) | 17213 (8.4) | 551502 (17.9) | 676098 (16.2) |
| 3 | 18775 (19.9) | 73994 (19.4) | 6532 (16.2) | 9085 (14.9) | 4933 (21.9) | 45595 (15.4) | 26627 (13.0) | 591164 (19.2) | 776705 (18.6) |
| 4 | 22558 (24.0) | 102260 (26.9) | 11466 (28.4) | 15083 (24.8) | 4395 (19.5) | 84321 (28.4) | 54357 (26.5) | 649457 (21.1) | 943897 (22.6) |
| 5 Most deprived | 21652 (23.0) | 109300 (28.7) | 14163 (35.0) | 26588 (43.7) | 2533 (11.2) | 128725 (43.3) | 91359 (44.5) | 754171 (24.5) | 1148491 (27.5) |
| Region of residence |  |  |  |  |  |  |  |  |  |
| North East | 2536 (2.7) | 6256 (1.6) | 259 (0.6) | 1996 (3.3) | 489 (2.2) | 5237 (1.8) | 2558 (1.2) | 174776 (5.7) | 194107 (4.6) |
| North West | 7935 (8.4) | 27196 (7.1) | 1344 (3.3) | 6846 (11.3) | 1368 (6.1) | 33309 (11.2) | 13710 (6.7) | 444311 (14.5) | 536019 (12.8) |
| Yorkshire &<br>Humber | 5516 (5.9) | 25459 (6.7) | 984 (2.4) | 6334 (10.4) | 1241 (5.5) | 29842 (10.0) | 9683 (4.7) | 338056 (11.0) | 417115 (10.0) |
| East Midlands | 4386 (4.7) | 28071 (7.4) | 1195 (3.0) | 3344 (5.5) | 1080 (4.8) | 16275 (5.5) | 10469 (5.1) | 277173 (9.0) | 341993 (8.2) |
| West Midlands | 6245 (6.6) | 26517 (7.0) | 2921 (7.2) | 6680 (11.0) | 1002 (4.4) | 42195 (14.2) | 14998 (7.3) | 339334 (11.0) | 439892 (10.5) |
| East of England | 8992 (9.5) | 43473 (11.4) | 2710 (6.7) | 3123 (5.1) | 2497 (11.1) | 20429 (6.9) | 15115 (7.4) | 346368 (11.3) | 442707 (10.6) |
| London | 35788 (38.0) | 138083 (36.3) | 24223 (59.9) | 25025 (41.1) | 8836 (39.2) | 111196 (37.4) | 107417 (52.3) | 337857 (11.0) | 788425 (18.9) |
| South East | 16023 (17.0) | 58048 (15.3) | 4640 (11.5) | 5338 (8.8) | 4301 (19.1) | 31634 (10.7) | 23331 (11.4) | 499489 (16.3) | 642804 (15.4) |
| South West | 6737 (7.2) | 27401 (7.2) | 2137 (5.3) | 2134 (3.5) | 1720 (7.6) | 6859 (2.3) | 8141 (4.0) | 316405 (10.3) | 371534 (8.9) |
| Maternal ethnic group |  |  |  |  |  |  |  |  |  |
| Bangladeshi | 58 (0.1) | 69 (<0.1) | 8 (<0.1) | 113 (0.2) | 10 (0.0) | 36575 (12.3) | 109 (0.1) | 10023 (0.3) | 46965 (1.1) |
| Indian | 616 (0.7) | 425 (0.1) | 135 (0.3) | 740 (1.2) | 281 (1.2) | 64014 (21.6) | 4045 (2.0) | 36985 (1.2) | 107241 (2.6) |
| Pakistani | 244 (0.3) | 1001 (0.3) | 14 (<0.1) | 1446 (2.4) | 113 (0.5) | 86836 (29.2) | 609 (0.3) | 49968 (1.6) | 140231 (3.4) |
| Black African | 132 (0.1) | 1364 (0.4) | 543 (1.3) | 3555 (5.8) | 110 (0.5) | 376 (0.1) | 89801 (43.7) | 8026 (0.3) | 103907 (2.5) |
| Black Caribbean | 44 (<0.1) | 193 (0.1) | 9629 (23.8) | 50 (0.1) | 111 (0.5) | 48 (<0.1) | 1669 (0.8) | 14863 (0.5) | 26607 (0.6) |
| White British | 9040 (9.6) | 35577 (9.3) | 1866 (4.6) | 2142 (3.5) | 5336 (23.7) | 2040 (0.7) | 11747 (5.7) | 2232271 (72.6) | 2300019 (55.1) |
| White Other | 9461 (10.0) | 212598 (55.9) | 7393 (18.3) | 6443 (10.6) | 8761 (38.9) | 1643 (0.6) | 10783 (5.2) | 56579 (1.8) | 313661 (7.5) |
| Other | 50530 (53.7) | 33285 (8.7) | 9375 (23.2) | 30646 (50.4) | 1803 (8.0) | 43852 (14.8) | 33258 (16.2) | 67878 (2.2) | 270627 (6.5) |
| Not known or missing | 24033 (25.5) | 95992 (25.2) | 11450 (28.3) | 15685 (25.8) | 6009 (26.7) | 61592 (20.7) | 53401 (26.0) | 597176 (19.4) | 865338 (20.7) |
| Maternal age |  |  |  |  |  |  |  |  |  |
| <20 | 866 (0.9) | 9797 (2.6) | 1195 (3.0) | 769 (1.3) | 210 (0.9) | 2153 (0.7) | 3220 (1.6) | 192498 (6.3) | 210708 (5.0) |
| 20-24 | 6509 (6.9) | 56946 (15.0) | 4483 (11.1) | 8899 (14.6) | 1300 (5.8) | 48360 (16.3) | 22830 (11.1) | 615649 (20.0) | 764976 (18.3) |
| 25-29 | 19486 (20.7) | 121150 (31.8) | 8915 (22.1) | 19568 (32.2) | 4399 (19.5) | 108930 (36.7) | 56274 (27.4) | 829993 (27.0) | 1168715 (28.0) |
| 30-34 | 34378 (36.5) | 121917 (32.0) | 13364 (33.1) | 18356 (30.2) | 8648 (38.4) | 92512 (31.2) | 69642 (33.9) | 847889 (27.6) | 1206706 (28.9) |
| 35-39 | 26352 (28.0) | 58812 (15.5) | 9619 (23.8) | 10245 (16.8) | 6228 (27.6) | 37470 (12.6) | 41461 (20.2) | 472364 (15.4) | 662551 (15.9) |
| 40-44 | 6177 (6.6) | 11302 (3.0) | 2664 (6.6) | 2798 (4.6) | 1639 (7.3) | 7043 (2.4) | 10912 (5.3) | 109565 (3.6) | 152100 (3.6) |
| 45+ | 390 (0.4) | 580 (0.2) | 173 (0.4) | 185 (0.3) | 110 (0.5) | 508 (0.2) | 1081 (0.5) | 5809 (0.2) | 8836 (0.2) |
| Missing | 0 (0.0) | 0 (0.0) | 0 (0.0) | 0 (0.0) | 0 (0.0) | 0 (0.0) | 2 (<0.01) | 2 (<0.01) | 4 (<0.01) |
| Sex |  |  |  |  |  |  |  |  |  |
| Female | 45274 (48.1) | 184639 (48.5) | 19675 (48.7) | 29339 (48.2) | 11008 (48.9) | 145337 (48.9) | 100651 (49.0) | 1496107 (48.7) | 2032030 (48.7) |
| Male | 48884 (51.9) | 195865 (51.5) | 20738 (51.3) | 31481 (51.8) | 11526 (51.1) | 151639 (51.1) | 104771 (51.0) | 1577662 (51.3) | 2142566 (51.3) |
| Congenital anomaly |  |  |  |  |  |  |  |  |  |
| No | 92292 (98.0) | 372235 (97.8) | 39590 (98.0) | 59289 (97.5) | 22029 (97.8) | 288759 (97.2) | 200488 (97.6) | 2991809 (97.3) | 4066491 (97.4) |
| Yes | 1866 (2.0) | 8269 (2.2) | 823 (2.0) | 1531 (2.5) | 505 (2.2) | 8217 (2.8) | 4934 (2.4) | 81960 (2.7) | 108105 (2.6) |

IMD = Index of multiple deprivation, SP = second parent

### Emergency admissions

76.0% (2,119,015/2,787,445) of hospital admissions in the cohort were emergency admissions (Table 4). Within world regional groups, unadjusted rates of emergency admissions were highest among children with UK-born mothers (171.6 per 1000 child-years, 95% CI 171.4-171.9), followed by children with mothers born in South Asia (155.9 per 100, 95% CI 155.1-156.7) and the Middle East and North Africa (129.1 per 100, 95% CI 127.5-130.8). Children whose mothers were born in Pakistan had the highest unadjusted emergency admission rates of the six maternal countries of birth studied (186.8 per 100, 95% CI 185.4-188.2), followed by mothers born in the UK and Bangladesh. Children with UK-born mothers and no second parent recorded on their birth registration had the highest emergency admission rates of all studied groups (219.7, 95% CI 218.5-220.9, Table 4).

**Table 4.** Observed numbers (%) and rates (per 1000 child-years) of emergency and planned hospital admissions, by maternal region of birth, maternal country of birth, migration status of parents and IMD group: England, births from 2008 to 2014.

|  | <u>Emergency admissions</u> |  | <u>Planned admissions</u> |  |
| --- | --- | --- | --- | --- |
|  | N (% of cases) | Rate per 1000 child-years (95% CI) | N (% of cases) | Rate per 1000 child-years (95% CI) |
| Overall | 2119015 (100.0) | 160.0 (159.8, 160.2) | 668430 (100.0) | 50.5 (50.3, 50.6) |
| **Maternal region of birth** |  |  |  |  |
| East-Asia & Pacific | 34343 (1.6) | 113.3 (112.1, 114.5) | 13228 (2.0) | 43.6 (42.9, 44.4) |
| Europe & Central Asia | 130673 (6.2) | 114.8 (114.2, 115.5) | 48957 (7.3) | 43.0 (42.6, 43.4) |
| Latin America & Caribbean | 14750 (0.7) | 114.3 (112.5, 116.2) | 6565 (1.0) | 50.9 (49.7, 52.1) |
| Middle East & North Africa | 24485 (1.2) | 129.1 (127.5, 130.8) | 11113 (1.7) | 58.6 (57.5, 59.7) |
| North America | 7120 (0.3) | 100.5 (98.2, 102.8) | 2867 (0.4) | 40.5 (39.0, 42.0) |
| South Asia | 145972 (6.9) | 155.9 (155.1, 156.7) | 53986 (8.1) | 57.7 (57.2, 58.1) |
| Sub-Saharan Africa | 77756 (3.7) | 116.7 (115.9, 117.5) | 33184 (5.0) | 49.8 (49.3, 50.3) |
| UK | 1683916 (79.5) | 171.6 (171.4, 171.9) | 498530 (74.6) | 50.8 (50.7, 51.0) |
| **Maternal country of birth** |  |  |  |  |
| Bangladesh | 24351 (1.1) | 148.9 (147.1, 150.8) | 9516 (1.4) | 58.2 (57.0, 59.4) |
| India | 34530 (1.6) | 129.2 (127.9, 130.6) | 11109 (1.7) | 41.6 (40.8, 42.4) |
| Nigeria | 14678 (0.7) | 108.6 (106.9, 110.4) | 6593 (1.0) | 48.8 (47.6, 50.0) |
| Pakistan | 68460 (3.2) | 186.8 (185.4, 188.2) | 25674 (3.8) | 70.0 (69.2, 70.9) |
| Poland | 39143 (1.8) | 104.8 (103.7, 105.8) | 15007 (2.2) | 40.2 (39.5, 40.8) |
| **Migration status of parents** |  |  |  |  |
| Both UK-born | 1426549 (67.3) | 169.1 (168.8, 169.4) | 421773 (63.1) | 50.0 (49.9, 50.2) |
| Mother UK-born & SP non-UK-born | 122092 (5.8) | 160.5 (159.6, 161.4) | 40899 (6.1) | 53.8 (53.2, 54.3) |
| Mother UK-born (sole registration) | 135275 (6.4) | 219.7 (218.5, 220.9) | 35858 (5.4) | 58.2 (57.6, 58.8) |
| Both non-UK-born | 300584 (14.2) | 123.2 (122.8, 123.6) | 119615 (17.9) | 49.0 (48.7, 49.3) |
| Mother non-UK-born & SP UK-born | 115676 (5.5) | 136.2 (135.4, 137.0) | 43070 (6.4) | 50.7 (50.2, 51.2) |
| Mother non-UK-born (sole registration) | 18839 (0.9) | 130.9 (129.0, 132.7) | 7215 (1.1) | 50.1 (49.0, 51.3) |
| **IMD groups** |  |  |  |  |
| 1 Least deprived | 278095 (13.1) | 137.8 (137.3, 138.3) | 93767 (14.0) | 46.5 (46.2, 46.8) |
| 2 | 315591 (14.9) | 147.9 (147.4, 148.5) | 101094 (15.1) | 47.4 (47.1, 47.7) |
| 3 | 374054 (17.7) | 152.5 (152.0, 152.9) | 119090 (17.8) | 48.5 (48.3, 48.8) |
| 4 | 479611 (22.6) | 160.6 (160.2, 161.1) | 151424 (22.7) | 50.7 (50.5, 51.0) |
| 5 Most deprived | 671664 (31.7) | 183.8 (183.4, 184.3) | 203055 (30.4) | 55.6 (55.3, 55.8) |

CI = Confidence interval, IMD = index of multiple deprivation, SP = second parent

Incidence rates (estimated from negative binomial regression models) of emergency admissions rose with increasing deprivation, but were most pronounced among children with mothers born in the UK, South Asia, and the Middle East and North Africa (30-45% increased rate comparing most with least deprived IMD group; Supplementary Table 7, Figure 3). Estimated rates for children with mothers born in Pakistan rose by IMD group in a pattern similar to that of children with UK-born mothers (Figure 3, Supplementary Table 8). Nigeria-born mothers were the only country of birth group to display no pattern by IMD group. Amongst parental migrant groups, children with both parents born outside the UK had the lowest estimated rates of admissions across all IMD groups (Figure 3).

**Figure 3.**
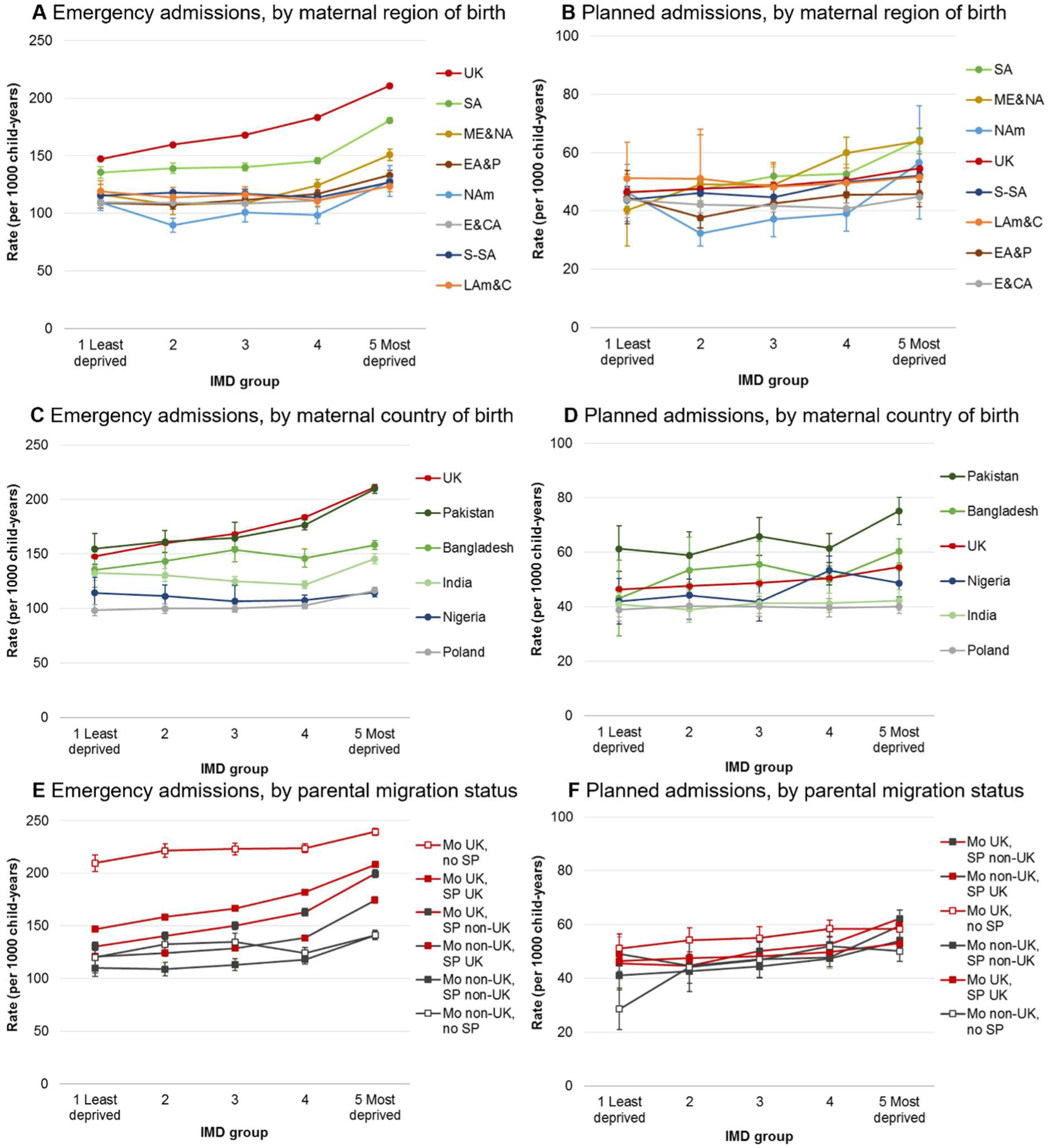
Estimated incidence rates of emergency and planned hospital admissions per 1000 person-years, by maternal world region of birth/maternal country of birth/parental migration status and IMD group (see Supplementary Tables 7-9 for further details, note Y-axis scales are different); IMD= index of multiple deprivation; EA&P= East Asia and Pacific, E&CA= Europe (excl. UK) and Central Asia, LAm&C= Latin America and Caribbean, ME&NA= Middle East and North Africa, NAm= North America, SA= South Asia, S-SA= Sub-Saharan Africa; Mo = mother, SP = second parent, UK = UK-born parent, Non-UK=non-UK-born parent

### Planned admissions

Within maternal region of birth groups, unadjusted rates of planned admissions were highest in children with mothers born in Middle East and North Africa (58.6 per 1000 child-years, 95% CI 57.5-59.7) and South Asia (57.7, 95% CI 57.2-58.1), Table 4. Children with mothers born in Pakistan and Bangladesh had unadjusted rates of planned admissions of 70.0 per 1000 child-years (95% CI 69.2-70.9) and 58.2 per 1000 (95% CI 57.0-59.4), respectively. Within parental migration status groups, children with UK-born mothers and no second parent registered had the highest rates of planned admissions (58.2 per 1000 child-years, 95% CI 57.6-58.8, Table 4). Rates of planned admissions estimated from negative binomial regression models broadly rose with increasing deprivation, but were most pronounced among children with mothers born in the Middle-East and North Africa (IRR 1.59, 95% CI 1.32-1.90, comparing most to least deprived IMD group) and South Asia (IRR 1.38, 95% CI 1.23-1.56), Supplementary Table 7, Figure 3.

### Secondary outcomes

Acute infections comprised 37.6% of all emergency admissions in this study (795,867/2,787,445), with an overall rate of 60.1 per 1000 child-years (95% CI 60.0-60.2; Table 5). Unadjusted and estimated rates of admissions for acute infections closely follow the patterns described for emergency admissions (Supplementary Figures 2-4, Supplementary Tables 10-12).

**Table 5.**
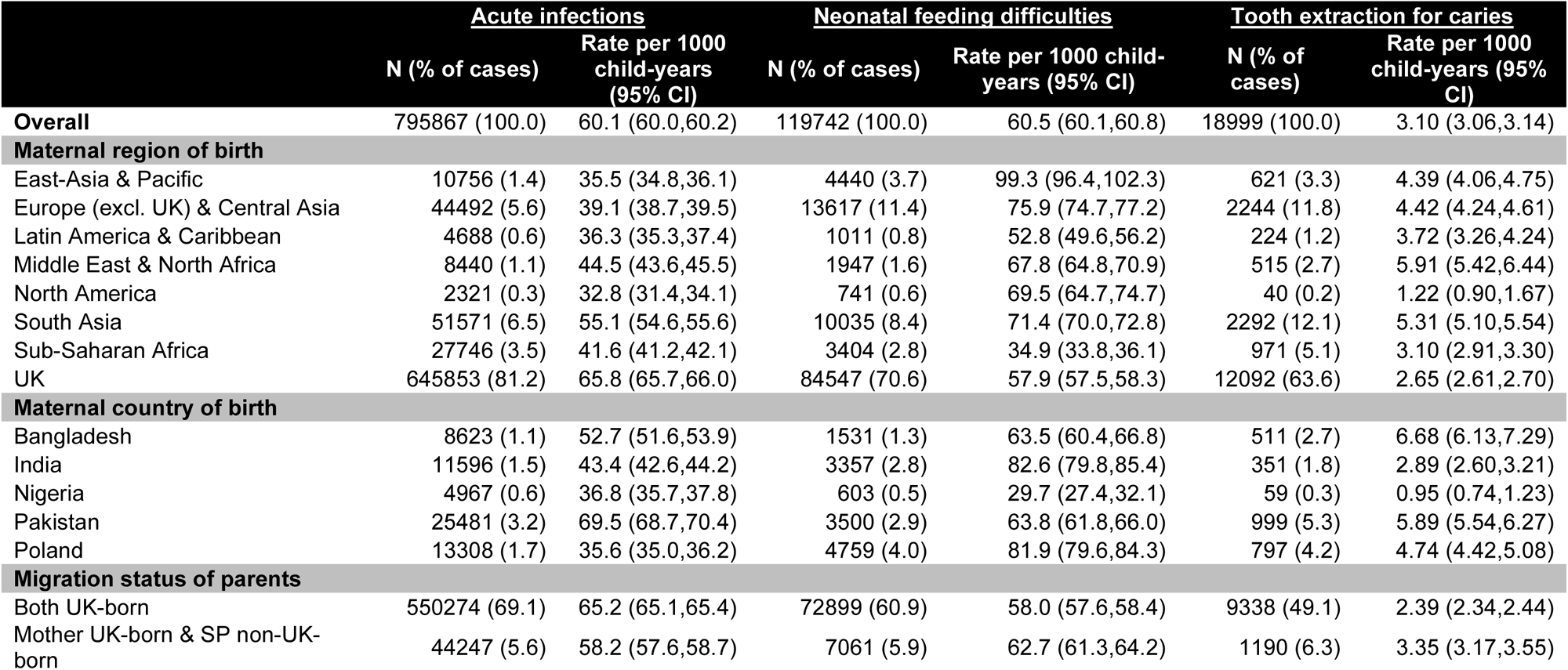
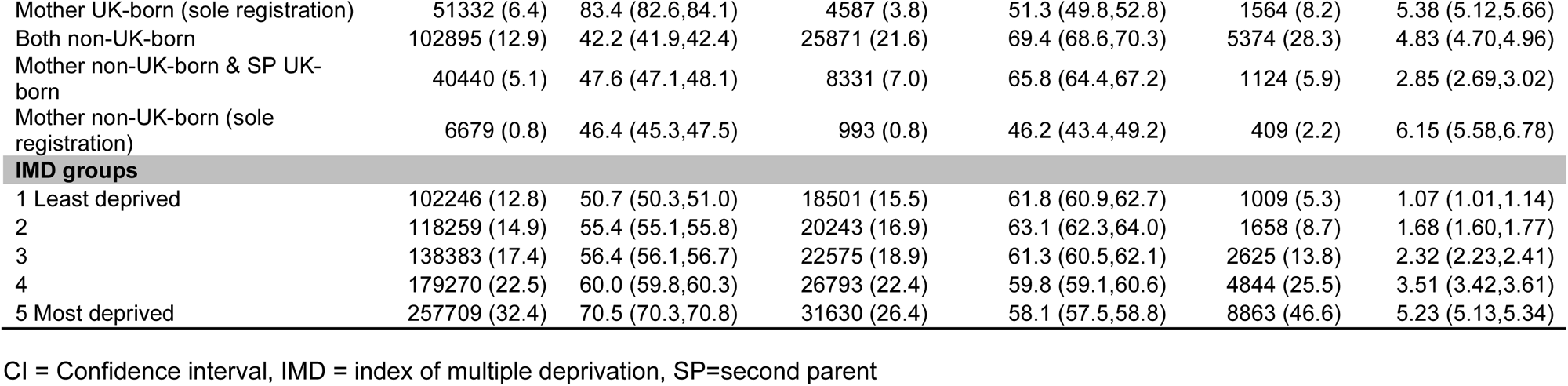
Observed numbers and rates (per 1000 child-years) of admissions with specified diagnoses, by maternal region of birth, maternal country of birth, migration status of parents and IMD group: England, births from 2008 to 2014.

There were 119,742 emergency admissions for feeding difficulties and jaundice in the first 6 months of life (4.3% of all emergency admissions), a rate of 60.5 per 1000 child-years (95% CI 60.1-60.8, Table 5). Unadjusted rates of admissions were highest among children of mothers born in East Asia and Pacific (99.3 per 1000, 95% CI 96.4-102.3) and Europe (excluding the UK) and Central Asia (75.9 per 1000, 95% CI 74.7-77.2). Estimated rates were highest for infants of mothers born in East Asia and Pacific (and, to a lesser extent, Europe and Central Asia) in the two most deprived IMD groups (Supplementary Figure 2, Supplementary Table 10). Conversely, infants of mothers born in other world regions display higher estimated incidence rates in the least deprived compared to the most deprived groups.

There were 18,999 planned admissions for tooth extraction including a diagnosis of caries (2.8% of the 668,430 total planned admissions), yielding a rate of 3.10 per 1000 child-years (95% CI 3.06-3.15, Table 5). These admissions were highest among children of mothers born in Middle East and North Africa (5.91 per 1000, 95% CI 5.42-6.44) and South Asia (5.31 per 1000, 95% CI 5.10-5.54). A steady pattern of increasing rates of admissions by IMD group is shown across all maternal region of birth groups, but most pronounced among children of mothers born in the UK (IRR 5.21, 95% CI 4.82-5.62) and the Midde East and North Africa (IRR 5.10, 95% CI 2.33-11.16; Supplementary Figure 2, Supplementary Table 10).

### Sensitivity analyses

Estimated rates of emergency and planned hospital admissions stratified by London and non-London residence at birth show similar patterns to national results presented above (Supplementary Figure 5). There were higher rates of emergency admissions across all maternal regions of birth and IMD groups for children living outside London compared to within London, and less differentiation between the maternal groups with the highest estimated rates (UK and South Asia) and the other world regions of birth. Results by emigration scenarios are presented in Supplementary Figures 6 and 7. The broad patterns of emergency admission rates were similar under the various emigration projections. However, the high level of estimated rates of emergency admission among children of UK-born mothers were matched by children of South Asia-born mothers only where it is assumed that one tenth of the UK-born population with mothers born in South Asia emigrate each year.

## Discussion

We used a national birth cohort dataset from England, including data from over four million children, to examine admission rates among parental migration and socioeconomic groups. We found that overall, children whose parents are born abroad were less likely to be admitted to hospital in an emergency compared to children whose parents are both born in the UK. Children of mothers born in the UK and no second parent registered at birth had the highest risk of emergency admissions. A socioeconomic gradient in emergency admissions was present, and was most pronounced for children of mothers born in the UK and South Asia, followed by the Middle East and North Africa. Planned admissions followed a similar socioeconomic gradient and were highest among children with mothers born in Middle East and North Africa, and South Asia. The high rates of emergency and planned admission estimated in the South Asia group were driven by children of women born in Pakistan and, to a lesser extent, Bangladesh. Rates of admissions for feeding difficulties and jaundice were highest for infants of East Asia and Pacific-born mothers in the two most deprived IMD groups, but highest in the least deprived groups among infants with mothers from Sub-Saharan Africa, Latin America and the Caribbean, and the UK. Admissions for tooth extractions due to caries displayed the greatest socioeconomic gradient, which was most pronounced among children of mothers born in the UK and the Middle East and North Africa.

### Strengths and limitations

To our knowledge, this is the first national UK-based study to examine hospital admission rates according to parental migration in England. Linkage between birth registration and hospital admission datasets allowed us to look at parental place of birth, which is not routinely collected in NHS data, and is a novel aspect of this research. Our results offer a starting point for further investigations into health inequities experienced by specific migrant communities, but the geographical regions (and even countries) of births used in this study comprise mixed groups of migrant women with diverse backgrounds and experiences of healthcare.^25^ Unmeasured sources of heterogeneity among our non-UK born sample includes the length of time since migration, reason for migration and English proficiency.^22,26^ Health inequity research using country of birth is well established in other European countries, with cited benefits including the objectivity and stability of this measure over time.^27^ In contrast, ethnicity, which is commonly used in health inequity research in the UK, is affected by reliability and completeness in routine data (and is missing for more than 20% of maternal records in this study).^7,28^ However, whilst there are overlaps between geographical place of birth and ethnicity, these are not the same entities, and the results of this study do not account for parents born in the UK (and elsewhere) who are from racially minoritised groups. Surveys, such as Understanding Society or the ONS Longitudinal Study, could be used in to incorporate important contextual and demographic factors into future research to further understand how multiple identities intersect to impact health and healthcare use among young children.

There was no missing information for maternal region of birth in this study, showing the value of the linked birth registration records. However, 8.1% of birth registrations were excluded from this study as they did not link to an NHS record (due to a mixture of linkage error and opt-outs leading to excluded records in the HES dataset). Linkage rates improved over study years, highlighting the value of replicating this research in future years. This reproduction is also important within the context of increasingly restrictive access to the NHS (and, more broadly, increasing levels of hostility and xenophobia) experienced by many migrants living in England, particularly following the UK’s Immigration Act of 2014 and 2016 and withdrawal from the European Union in 2020.^29^ Only area-level measures of socioeconomic position were available in our administrative data sources, meaning that we are likely to have underestimated the true family-level effect of this determinant of health.^30^ IMD scores are weighted towards the income and employment domains, which are constructed using data on benefits, meaning that IMD as a whole may be less reliable as an indicator of disadvantage amongst migrant groups (due to lower take up of benefits, on average).^31,32^ Replication of this analyses with a socioeconomic indicator measured at the family-level, such as household income or parental education, may help to improve estimation.

### Interpretation and implications

Unlike patterns reported in other high income countries,^6^ we found that emergency admission rates were generally higher for children of UK-born parents compared with non-UK born parents, even at the same or higher levels of socioeconomic deprivation. This finding matches results of a study of linked hospital-general practice registration records in England, which estimated that recent migrants to England (indicated by first registration with a general practice after the age of 15 years) had about half the rate of hospital admissions compared with the general population of over 15 year olds.^33^ There is evidence to suggest that accident and emergency use in England is also lower amongst children of non-UK born mothers. Using hospital data linked to the Born in Bradford cohort study, Credé et al. estimated that the odds of accident and emergency use in the first five years of life was 0.92 times lower among children of mothers from UK/Ireland compared to children of mothers from other countries (OR 0.92, 95% CI 0.83, 1.00).^22^ Collectively, these findings help to dispel a common (but unevidenced) perception that migrants and their young children place excessive strain on the UK NHS.^34^ Two contrasting explanations can be posited for this major finding: that these young children are healthier and therefore require less healthcare interventions (a form of the “healthy migrant effect”);^35^ or that migrant parents and their children face barriers in accessing timely and preventative health services, including emergency care where needed.^22^ A third explanation, of returning home for hospital treatment, as has been reported by migrants from Eastern Europe,^36^ may alternatively explain particularly low admission rates for some groups of children.

The healthy migrant effect is the phenomenon, whereby, even after accounting for selection effects, migrants show similar or better health (for some outcomes) than the non-migrant population despite higher levels of socioeconomic deprivation.^35^ Factors that may contribute to resilience in specific communities, and warrant further investigation, include the buffering effects of living in areas with other migrants and minoritised groups who may provide support and signposting to preventive care,^37^ and the role of culturally-specific service provision (a factor that emerged from patient and public involvement undertaken to inform this study). However, beyond the headline finding, we find divergent patterns of hospital admissions by maternal regions/countries of birth, which suggest a more complex pattern of secondary healthcare use by geographical origin of birth. This reflects quantitative and qualitative studies of the perinatal period, which report mixed experiences of healthcare and child health outcomes in the UK across migrant women.^38,39^ Evidence of poorer early childhood health for children of mothers born in South Asia, particularly Pakistan, including increased risk of low birthweight, preterm birth and congenital anomalies,^40,41^ fits with the relatively high emergency and planned hospital admission rates shown here. However, poor early life outcomes are also reported in the UK for women born in Africa, including high rates of infant mortality,^42^ sits in contrast to the relatively low emergency hospital admission rates for children of mothers born in Sub-Saharan Africa in this study. This is particularly striking given that almost half of children with mothers born in Sub-Saharan Africa were in the most deprived socioeconomic group. Healthcare barriers specific to some group may include: less culturally and linguistically appropriate services; reduced awareness of healthcare entitlements; or, particularly in vulnerable groups, a fear of charging, or immigration enforcement.^43,44^ Linkage between birth registration and/or Census data to primary care and accident and emergency attendances, as well as outpatient datasets, and qualitative studies within diverse groups of parents who have migrated to the UK will help to elucidate whether health service utilisation is proportionate to need. Future research could focus on children with specific conditions, such as asthma or epilepsy, to capture differences in care pathways across children from different backgrounds.

A socioeconomic gradient in the rates of hospital admissions in England, as shown in this study, has been repeatedly documented in the general childhood population.^45,46^ Emergency hospital use is associated with poorer access to primary care and lower engagement with preventative services, and rates of both emergency and planned admissions are higher amongst children with chronic conditions (the prevalence of which is also socially graded).^46^ Planned admissions for tooth extractions for caries clearly reflects substantial oral health inequalities,^47^ whereas admissions for feeding difficulties and jaundice illustrate a more complex socioeconomic pattern of secondary healthcare use. Exclusive breastfeeding (which is linked to the onset of feeding difficulties and physiological jaundice) is more common amongst women from affluent backgrounds in England, which likely explains the reverse social gradient shown for these admissions.^48^ However, the reverse pattern amongst mothers born in East Asia and Pacific highlights that this inference may not be generalisable to all women.

On the whole, this evidence suggests that a continued focus on improving access to preventative health services, including antenatal, primary care and dental services, in poorer areas is necessary for all children, but with consideration of the diverse needs of parents and children from different migrant backgrounds. Support for children whose births are not jointly registered (a known vulnerable group of families) is also warranted given the high rates of admissions across all socioeconomic strata of this group.^49^ In addition, strengthening public health programmes such as diet, overcrowding and environmental tobacco smoke, is a complementary pathway to improving the health of all children. This should be a focus for Integrated Care Systems and the renewal of the Healthy Child Programme in England.^50^

## Conclusion

This research indicates that children whose parents who have migrated to the UK generally have lower overall usage of NHS emergency secondary care services than children of UK-born parents. Our study revealed a socioeconomically graded patterns of hospital admissions for all children born in England, which were highest amongst those with mothers born in the UK, South Asia, and the Middle East and North Africa. Future research using linked primary and secondary care datasets will improve understanding on whether healthcare use is proportionate to need.

## Data Availability

The authors do not have permission to supply data or identifiable information to third parties. Anyone wishing to access the linked datasets for research purposes should apply via the CAG to the Health Research Authority to access patient-identifiable data without consent and then to the ONS and NHS Digital. In the first instance, enquiries about access to the data analysed here should be addressed to the corresponding author and enquiries about the City Birth Cohort should be addressed to Alison Macfarlane.

## Authors’ contributions

All authors have contributed to this study. KML planned and designed the study based on an initial idea and conception by PH. KML conducted statistical analysis with support from PH & MCB. KML prepared the first manuscript draft, and all authors provided substantial contribution to interpretation of the results and manuscript revisions. PH and RV obtained funding to conduct the study. All authors approved the final manuscript; and have read, and confirm that they meet, International Committee of Medical Journal Editors (ICMJE) criteria for authorship.

## Ethics

We have obtained ethical or information governance approvals from the following committees: NHS London Queen Square Ethics Committee (full approval, reference: 18/LO/1514); Confidentiality Advisory Group (full approval, reference: 18/CAG/0159); Administrative Data Research Network (approval, reference PROJ-194); ONS Research Accreditation Panel (full approval, reference 2019/020); National Statistician’s Data Ethics Advisory Committee (full approval, reference: 18 (07)); Independent Group Advising on Release of Data (NHS Digital; full approval, DARS-NIC-234656).

Approvals for the City Birth Cohort were obtained as follows. Ethics approval 05/Q0603/108 and subsequent substantial amendments were granted by East London and City Local Research Ethics Committee 1 and its successors. Permissions to use patient identifiable data without consent PIAG 2-10(g)/2005 and CAG 9-08(b) 2014 were granted by the Confidentiality Advisory Group and its predecessors. Permission to access data from the Office for National Statistics in the VML, now known as the Secure Research Service was granted by ONS’s Microdata Release Panel, now superseded by its Research Advisory Panel. Permission to link and analyse data held by the Health and Social Care Information Centre, now known as NHS Digital, was granted under Data Sharing Agreements NIC-273840-N0N0N and subsequently under DARS-NIC-10094-P6P4B-v6.7

## Disclaimers

This work uses data provided by patients and collected by the NHS as part of their care and support. The use of Hospital Episodes Statistics data was approved by NHS Digital for the purpose of this study. Source data can be accessed by researchers applying to the Health and Social Care Information Centre for England. Copyright © 2018. Reused with the permission of the Health and Social Care Information Centre. All rights reserved.

The views expressed in this publication are those of the author(s) and not necessarily those of the NHS, the National Institute for Health Research, the Department of Health and Social Care or its arm’s length bodies, and other Government Departments.

The work was done in the Secure Environment of ONS’ Secure Research Service. The use of the ONS statistical data in this work does not imply the endorsement of the ONS in relation to the interpretation or analysis of the statistical data. This work uses research datasets which may not exactly reproduce National Statistics aggregates.

## Abbreviations

AIC: Akaike Information Criterion
GP: General practitioner
HES APC: Hospital Episode Statistics Admitted Patient Care
ICD-10: International Classification of Diseases 10th Revision
IDACI: Income Deprivation Affecting Children Index
IRR: Incidence Rate Ratio
NHS: National Health Service
MCS: Millennium Cohort Study
ONS: Office for National Statistics

