## Supplementary file for "Parental migration, socioeconomic deprivation and hospital admissions in preschool children: national cohort study"

### Supplementary material

#### Appendix A. Defining study outcomes and covariates

**Supplementary Table 1. Diagnosis code list for secondary study outcomes (primary diagnoses and emergency admissions unless specified)**

| Condition(s) | ICD-10 name | ICD-10 code |
|---|---|---|
| **Acute infections - lower respiratory tract infections** | | |
| Influenza, | Influenza with pneumonia, virus identified | J10.0 |
| pneumonia | Influenza with pneumonia, virus not identified | J11.0 |
| | Influenza with other respiratory manifestations, virus not | J11.1 |
| | identified | J12 |
| | Viral pneumonia, not elsewhere classified | J13.X |
| | Pneumonia due to Streptococcus pneumoniae | J14 |
| | Pneumonia due to Haemophilus influenzae | J15 |
| | Bacterial pneumonia, not elsewhere classified | J16.0 |
| | Chlamydial pneumonia | J16.8 |
| | Pneumonia due to infectious organisms not elsewhere | J18.0 |
| | classified | J18.1 |
| | Bronchopneumonia, unspecified | J18.8 |
| | Lobar pneumonia, unspecified | J18.9 |
| | Other pneumonia, organism unspecified | |
| | Pneumonia, unspecified | |
| Bronchiolitis | Acute bronchiolitis | J21 |
| | Unspecified acute lower respiratory infection | J22 |
| **Acute infections - upper respiratory tract infections** | | |
| Otitis media | Suppurative and unspecified otitis media | H66 |
| | Otitis media in diseases classified elsewhere | H67 |
| Throat | Acute pharyngitis | J02 |
| infections | Acute tonsillitis | J03 |
| | Acute laryngitis | J04.0 |
| | Acute upper respiratory infections of multiple & unspecified | J06 |
| | sites | J31.2 |
| | Chronic pharyngitis | J05 |
| | Acute obstructive laryngitis [croup] and epiglottitis | |
| **Acute infections - urinary tract infections (or pyelonephritis)** | | |
| Kidney | Acute tubulo-interstitial nephritis | N10 |
| infections | Chronic tubulo-interstitial nephritis | N11 |
| | Tubulo-interstitial nephritis, not specified as acute or chronic | N12 |
| | Pyonephrosis | N13.6 |
| | Renal tubulo-interstitial disease, unspecified | N15.9 |
| Cystitis (bladder | Acute cystitis | N30.0 |
| inflammation) | Other cystitis | N30.8 |

| | | |
|---|---|---|
| | Cystitis, unspecified | N30.9 |
| | Urinary tract infection, site not specified | N39.0 |
| Dehydration and gastroenteritis | | |
| | Volume depletion | E86 |
| | Allergic and dietetic gastroenteritis and colitis | K52.2 |
| | Other specified noninfective gastroenteritis and colitis | K52.8 |
| | Noninfective gastroenteritis and colitis, unspecified | K52.9 |
| | Salmonella enteritis | A02.0 |
| | Other bacterial intestinal infections | A04 |
| | Bacterial foodborne intoxication, unspecified | A05.9 |
| | Cryptosporidiosis | A07.2 |
| | Viral and other specified intestinal infections | A08 |
| | Other gastroenteritis and colitis of infectious & unspecified origin | A09 |
| | Gastroenteritis and colitis due to radiation | K52.0 |
| | Toxic gastroenteritis and colitis | K52.1 |
| Other/unknown | | |
| | Viral infection of unspecified site | B34 |
| Feeding difficulties and jaundice | | |
| | Neonatal jaundice from other and unspecified causes | P59 |
| | Feeding problems of new-born | P92 |
| Tooth extractions for caries* (planned admissions only) | | |
| Dental caries | Caries of dentine | K02.1 |
| | Dental caries on pit and fissure surface | K02.5 |
| | Other dental caries | K02.8 |
| | Dental caries, unspecified | K02.9 |
| | Diseases of pulp and periapical tissues | K04.0 |
| | Chronic apical periodontitis | K04.5 |
| | Periapical abscess with sinus | K04.6 |
| | Periapical abscess without sinus | K04.7 |
| Tooth extraction | Surgical removal of tooth (main operative procedure) | F09 (OPCS code) |
| | Simple extraction of tooth (main operative procedure) | F10 (OPCS code) |

ICD-10 = International Classification of Diseases and Related Health Problems 10th Revision; OPCS = Office of Population Census and Survey codes (surgical procedure codes); *To meet definition, child must have any primary diagnosis of dental caries AND one of the two listed operative procedures (as outlined in the NHS Outcomes Framework indicator specification)[1]

##### *1.1. Covariates*

Child sex (female or male), geographical region of residence (London, the North East, North West, Yorkshire and the Humber, East Midlands, West Midlands, South East, East of England, the South West), year of birth (2008 to 2014), and maternal age at the birth of the child (split into <20, 20-29, 30-39, 40+ years) were drawn from ONS birth registration records. Parity, defined as the mother's number of previous live or stillbirths (split into 0, 1, 2+), was derived from ONS birth registrations and supplemented with linkage of HES APC delivery records if missing. Presence of congenital anomalies (yes or no), based on the Hardelid UK chronic condition ICD-10 code list,[4] were identified in infant hospital admissions or death certificates up to age two years.

**Supplementary Table 2. ICD-10 codes used to define congenital anomalies**

| ICD-10 subchapter | ICD-10 codes |
|---|---|
| Congenital malformation of the nervous system | Q00 Q01 Q02 Q03 Q04 Q05 Q06 Q07 |
| Congenital malformations of eye, ear, face and neck | Q104 Q107 Q11 Q12 Q130-Q134 Q138 Q139 Q14 Q15 Q16 Q188 |
| Congenital malformations of the circulatory system | Q20 Q21 Q22 Q23 Q24 Q25 Q26 Q27 Q28 |
| Congenital malformations of the respiratory system | Q30 Q31 Q32 Q33 Q34 |
| Cleft lip and cleft palate | Q35 Q36 Q37 |
| Other congenital malformations of the digestive system | Q380 Q383 Q384 Q386-Q388 Q39 Q402-Q409 Q41 Q42 Q431 Q433-Q437 Q439 Q44 Q45 |
| Congenital malformations of genital organs | Q500 Q51 Q520-Q522 Q524 Q540-Q543 Q548 Q549 Q550 Q555 Q56 |
| Congenital malformations of the urinary system | Q601 Q602 Q604-Q606 Q61 Q620-Q626 Q628 Q630-Q632 Q638 Q639 Q64 |
| Congenital malformations and deformations of the musculoskeletal system | Q650-Q652 Q658 Q659 Q675 Q682-Q685 Q71 Q72 Q73 Q74 Q750 Q751 Q753-Q759 Q761-Q764 Q77 Q78 Q790 Q792-Q798 |
| Other congenital malformations | Q80 Q81 Q820-Q824 Q829 Q85 Q86 Q87 Q891-Q899 |
| Chromosomal abnormalities not elsewhere classified | Q90 Q91 Q92 Q93 Q952-Q953 Q97 Q980 Q99 |

ICD-10 = International Classification of Diseases and Related Health Problems 10th Revision

#### 2. Appendix B. Theoretical diagram and confounder selection

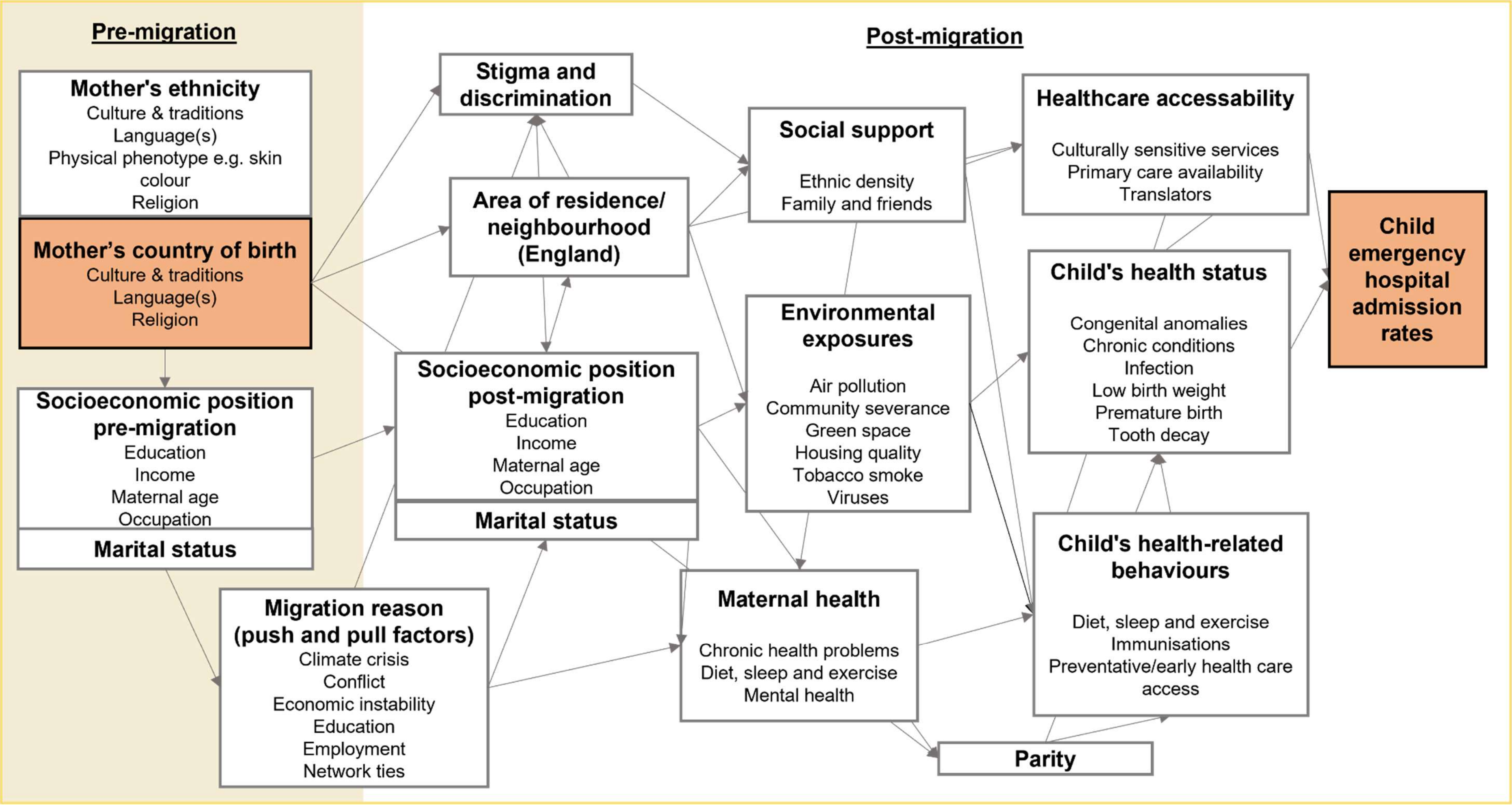

**Supplementary Figure 1. DAG showing potential pathways between maternal country of birth and offspring emergency hospital admissions rates; Orange box indicates exposure or outcome**

#### 3. Appendix C. Sensitivity analyses - estimating emigration in the national birth cohort

##### *3.1. Outlining the problem*

Our study datasets do not include information on emigration of the children in the cohort during follow up. Not including this information may bias the results by overestimating the denominator (i.e. the person-years) and therefore underestimating rates of admissions among emigrants. Given that some populations are more likely to be mobile, the impact of this bias is unlikely to be uniform across maternal world region of birth or parental migration status groups. It is therefore important to carry out some sensitivity analyses assuming different scenarios of emigration to assess how this bias may affect the results of our study. We do not need to measure immigration or net migration for our work because our cohort includes all children born in England to resident women.

##### *3.2. Suggested solutions*

The ideal solution to our problem would be individual-level linkage to records of emigration. This is not currently possible. We are due to get embarkation dates from the Personal Demographics Service (PDS) for the children in the cohort from NHS Digital. The PDS will include records of anyone who has let their GP or other NHS care provider know they are moving out of England. However, these data are likely to be highly incomplete, as not all those who emigrate alert their GP. The second best option is to impute probable rates of emigration based on aggregate information of rates of emigration, matched on key variables such as maternal age and region of birth. This requires a dataset(s) with a reliable measure of the size (or stock) of the population at risk of migration and emigration rates over time.

##### *3.3. Official statistics*

Estimating migration stock: the Annual Population Survey (APS) provides an estimate of the number of people living in private households in the UK, by geographical area and key characteristics (including country of birth).[7] The APS is created by combining 2 waves of the main Labour Force Survey (a continuous household survey of the UK) with data collected on a local sample boost.

Estimating migration flows: The official source of data on immigration and emigration, produced by the Office for National Statistics (ONS), is the Long-Term International Migration (LTIM) series (previously called the Total International Migration statistics or TIM).[8] LTIM is primarily created using information from the International Passenger Survey (IPS), which is a sample survey carried out at all main ports in the UK to capture migration intentions.[9] Owing to some methodological problems with the IPS (including sampling problems and uncertain intentions by survey respondents), the data since 2008 has been adjusted using other data sources. Since 2019, the LTIM is classified as experimental statistics whilst further work is being undertaken to improve this dataset.

##### *3.4. Conclusion from available data*

Theoretically it would be possible to combine data from LTIM and the APS to calculate migration rates according to sex, age and region of birth, which would allow us to carry out the above imputation analyses. However, the ONS has specifically written that the LTIM and APS dataset should not be used in conjunction with one another to estimate the proportion of the population who migrate.[5] As such, there is no one single source of data in the UK that can be used to reliably model the proportion of the population that emigrates over a period of time. In any case, neither the LTIM or the APS have data specifically on the emigration of women who have given birth in the last five years, nor migration of children aged less than five years old according to their parents' country of birth. The ONS is in the process of transforming population and migration statistics by using a combination of administrative data sources and has written about producing triangulated population stock and flow data by population characteristics.[10] But this information is not yet available, and may not cover our study years (2008 to 2014) in any case.

##### *3.5. Working solution*

In the absence of reliable data on emigration, we will therefore model four simple scenarios, each with different levels of assumed annual emigration by maternal world region of birth (as shown in Supplementary Table 3). The first two scenarios consider all groups apart from the UK-born group to have the same (higher) levels of emigration. The last two scenarios consider higher levels of emigration among children with mothers born in East-Asia and Pacific, Europe (excluding the UK) & Central Asia, and North America. This differentiation is based on a Home Office report on patterns of long term emigration in the UK up to 2011, which concludes that people from higher-income non-UK countries (including EU countries, New-Zealand and Australia) are less likely to stay permanently in the UK.[11]

We make several additional assumptions when doing these simulations:

1. For each age group each year a uniform percentage of children emigrate
2. Emigration implies not returning to England during follow up time
3. Children who migrate do so half-way through the year of follow up

Supplementary table 4 outlines how the data were transformed in each emigration scenario. Following this process, the data were analysed according to the methods described in the main paper (for maternal region of birth as the exposure and emergency admissions rates as the outcome).

**Supplementary Table 3. Assumed levels of emigration (*x* in supplementary table 4) for each scenario, by maternal world region of birth**

| | Scenario 1 | Scenario 2 | Scenario 3 | Scenario 4 |
|---|---|---|---|---|
| East-Asia & Pacific | 5% | 10% | 5% | 10% |
| Europe (excl. UK) & Central Asia | 5% | 10% | 5% | 10% |
| Latin America & Caribbean | 5% | 10% | 1% | 1% |
| Middle East & North Africa | 5% | 10% | 1% | 1% |
| North America | 5% | 10% | 5% | 10% |
| South Asia | 5% | 10% | 1% | 1% |
| Sub-Saharan Africa | 5% | 10% | 1% | 1% |
| UK | 1% | 1% | 1% | 1% |

**Supplementary Table 4. Method to apply emigration projections within maternal region of birth groups**

| Year | Child age (years) | Method (where *x* = % specified in Supplementary Table 2) |
|---|---|---|
| 2008 | 0 | Set end of follow up to 0.5 years for random x% of cohort members (only among those with no admissions in follow up) |

| | | |
|---|---|---|
| 2009 | 0, 1 | Set end of follow up to 0.5 (for those born in 2009) or 1.5 (for those born in 2008) years for random x% of cohort members within each birth year (among those with no cases between 2009 and 2014) |
| 2010 | 0, 1, 2 | Set end of follow up to 0.5 (for those born in 2010), 1.5 (for those born in 2009) or 2.5 (for those born in 2008) years for random x% of cohort members within each birth year (among those with no cases between 2010 and 2014) |
| 2011 | 0, 1, 2, 3 | Set end of follow up to 0.5 (for those born in 2011), 1.5 (for those born in 2010), 2.5 (for those born in 2009) or 3.5 (for those born in 2008) years for random x% of cohort members within each birth year (only those with no cases between 2011 and 2014) |
| 2012 | 0, 1, 2, 3, 4 | Set end of follow up to 0.5 (for those born in 2012), 1.5 (for those born in 2011), 2.5 (for those born in 2010), 3.5 (for those born in 2009) or 4.5 (for those born in 2008) years for random x % of cohort members within each birth year (only those with no cases between 2012 and 2014) |
| 2013 | 0, 1, 2, 3, 4 | Set end of follow up to 0.5 (for those born in 2013), 1.5 (for those born in 2012), 2.5 (for those born in 2011), 3.5 (for those born in 2010) or 4.5 (for those born in 2009) years for random x% of cohort members within each birth year (only those with no cases between 2013 and 2014) |
| 2014 | 0, 1, 2, 3, 4 | Set end of follow up to 0.5 (for those born in 2014), 1.5 (for those born in 2013), 2.5 (for those born in 2012), 3.5 (for those born in 2011) or 4.5 (for those born in 2010) years for random x% of cohort members within each birth year (only those with no cases in 2014) |

**Supplementary Table 5. Key characteristics of all children (pre-linkage) and children without ONS-HES APC record linkage**

| | Total | Missing HES APC record | | Association with missing HES APC record* |
|---|---|---|---|---|
| | N | N | % | OR (95% CI) |
| Overall | 4,560,665 | 371,316 | 8.1 | |
| Maternal region of birth | | | | |
| East-Asia & Pacific | 101,727 | 7,280 | 7.2 | 1.26 (1.23, 1.30) |
| Europe & Central Asia | 409,952 | 28,164 | 6.9 | 1.21 (1.19, 1.23) |
| Latin America & Caribbean | 44,720 | 4,110 | 9.2 | 1.66 (1.60, 1.72) |
| Middle East & North Africa | 65,460 | 4,404 | 6.7 | 1.18 (1.14, 1.22) |
| North America | 25,024 | 2,409 | 9.6 | 1.75 (1.67, 1.83) |
| South Asia | 316,490 | 18,203 | 5.8 | Ref. |
| Sub-Saharan Africa | 227,172 | 20,749 | 9.1 | 1.65 (1.61, 1.68) |
| UK | 3,369,437 | 285,314 | 8.5 | 1.52 (1.49, 1.54) |
| Missing | 683 | 100 | 14.6 | |
| Migration status of parents | | | | |
| Both UK-born | 2,891,186 | 234,689 | 8.1 | 1.24 (1.23, 1.25) |
| Mother UK-born & SP non-UK-born | 261,888 | 23,431 | 8.9 | 1.38 (1.36, 1.40) |
| Mother UK-born (sole registration) | 216,363 | 27,194 | 12.6 | 2.02 (1.98, 2.05) |
| Both non-UK-born | 848,492 | 56,497 | 6.7 | Ref. |
| Mother non-UK-born & SP UK-born | 291,820 | 23,808 | 8.2 | 1.25 (1.23, 1.27) |
| Mother non-UK-born (sole registration) | 50,766 | 5,048 | 9.9 | 1.55 (1.50, 1.60) |
| Missing | 150 | 66 | 44.0 | |
| IMD groups | | | | |
| 1 Least deprived | 636,622 | 49,641 | 7.8 | Ref. |
| 2 | 748,473 | 58,748 | 7.8 | 1.01 (0.99, 1.02) |
| 3 | 877,155 | 71,143 | 8.1 | 1.04 (1.03, 1.06) |
| 4 | 1,038,197 | 82,818 | 8.0 | 1.03 (1.01, 1.04) |
| 5 Most deprived | 1,256,984 | 107,087 | 8.5 | 1.10 (1.09, 1.11) |
| Missing | 3,234 | 1,296 | 40.1 | |
| Year of birth | | | | |

| | | | | |
|---|---|---|---|---|
| 2008 | 648,609 | 96,615 | 14.9 | 2.70 (2.67, 2.73) |
| 2009 | 646,192 | 61,274 | 9.5 | 1.62 (1.60, 1.64) |
| 2010 | 658,283 | 48,035 | 7.3 | 1.21 (1.20, 1.23) |
| 2011 | 659,188 | 43,560 | 6.6 | 1.09 (1.08, 1.11) |
| 2012 | 669,505 | 43,008 | 6.4 | 1.06 (1.04, 1.07) |
| 2013 | 641,217 | 39,023 | 6.1 | Ref. |
| 2014 | 637,671 | 39,218 | 6.2 | 1.01 (1.00, 1.03) |
| Region of residence | | | | |
| North East | 202,401 | 7,615 | 3.8 | Ref. |
| North West | 595,580 | 57,592 | 9.7 | 2.74 (2.67, 2.81) |
| Yorkshire & Humber | 448,229 | 29,603 | 6.6 | 1.81 (1.76, 1.86) |
| East Midlands | 368,236 | 24,946 | 6.8 | 1.86 (1.81, 1.91) |
| West Midlands | 485,901 | 44,096 | 9.1 | 2.55 (2.49, 2.62) |
| East of England | 485,977 | 41,725 | 8.6 | 2.40 (2.34, 2.46) |
| London | 865,799 | 74,329 | 8.6 | 2.40 (2.35, 2.46) |
| South East | 706,711 | 61,792 | 8.7 | 2.45 (2.39, 2.51) |
| South West | 401,610 | 28,814 | 7.2 | 1.98 (1.93, 2.03) |
| Missing | 221 | 221 | 100.0 | |

OR=odds ratio, SP = second parent;*unadjusted logistic regression of the association between key characteristics and no available HES APC birth record (due to opt outs and non-linkage)

#### 4. Appendix D. Largest maternal countries of birth within each world region

**Supplementary Table 6. Five largest maternal countries of birth within each world region (defined by the number of children in the birth cohort)**

| East-Asia & Pacific (*N* = 94,158) | | | |
|---|---|---|---|
| | Country | N | % |
| 1 | China | 21,416 | 22.7 |
| 2 | Philippines | 16,672 | 17.7 |
| 3 | Australia | 12,583 | 13.4 |
| 4 | New Zealand | 6,693 | 7.1 |
| 5 | Thailand | 6,526 | 6.9 |

| North America (*N* = 22,534) | | | |
|---|---|---|---|
| | Country | N | % |
| 1 | United States | 15,620 | 69.3 |
| 2 | Canada | 6,600 | 29.3 |
| 3 | Bermuda | 310 | 1.4 |
| 4 | # | # | # |
| 5 | # | # | # |

| Europe & Central Asia (*N* = 380,504) | | | |
|---|---|---|---|
| | Country | N | % |
| 1 | Poland* | 122,889 | 32.3 |
| 2 | Germany | 29,050 | 7.6 |
| 3 | Romania | 23,438 | 6.2 |
| 4 | Lithuania | 23,209 | 6.1 |
| 5 | Ireland | 17,503 | 4.6 |

| South Asia (*N* = 296,976) | | | |
|---|---|---|---|
| | Country | N | % |
| 1 | Pakistan* | 115,886 | 39.0 |
| 2 | India* | 85,830 | 28.9 |
| 3 | Bangladesh* | 50,831 | 17.1 |
| 4 | Sri Lanka | 21,357 | 7.2 |
| 5 | Afghanistan | 17,495 | 5.9 |

| Latin America & Caribbean (*N* = 42,211) | | | |
|---|---|---|---|
| | Country | N | % |
| 1 | Jamaica | 12,728 | 31.5 |
| 2 | Brazil | 8,984 | 22.2 |
| 3 | Colombia | 3,217 | 8.0 |
| 4 | Trinidad and Tobago | 1,755 | 4.3 |
| 5 | Ecuador | 1,448 | 3.6 |

| Sub-Saharan Africa (*N* = 205,422) | | | |
|---|---|---|---|
| | Country | N | % |
| 1 | Nigeria* | 42,934 | 20.9 |
| 2 | Somalia | 33,614 | 16.4 |
| 3 | South Africa | 25,032 | 12.2 |
| 4 | Ghana | 20,735 | 10.1 |
| 5 | Zimbabwe | 16,189 | 7.9 |

| Middle East & North Africa (*N* = 60,820) | | | |
|---|---|---|---|
| | Country | N | % |
| 1 | Iraq | 13,988 | 23.0 |
| 2 | Algeria | 5,967 | 9.8 |
| 3 | Iran | 5,594 | 9.2 |
| 4 | Morocco | 4,603 | 7.6 |
| 5 | Libya | 4,519 | 7.4 |

| UK* (*N* = 3,073,769) | | | |
|---|---|---|---|
| | Nation/country | N | % |
| 1 | England | 2,987,370 | 97.2 |
| 2 | Scotland | 39,444 | 1.3 |
| 3 | Wales | 31,569 | 1.0 |
| 4 | Northern Ireland | 13,194 | 0.4 |
| 5 | Jersey | 845 | <0.01 |

### Supressed due to low cell count and other values rounded to nearest 10; *Included in country of birth analyses

#### 5. Appendix E. Results

**Supplementary Table 7. Estimated incidence rates and adjusted IRRs of hospital admissions, by maternal region of birth and IMD group (derived from negative binomial regression models)***

| | Emergency admissions** | | | Planned admissions*** | | |
|---|---|---|---|---|---|---|
| | Incidence rate (95% CI) per 1000 child-years | IRR (95% CI) | p-value | Incidence rate (95% CI) per 1000 child-years | IRR (95% CI) | p-value |
| East-Asia & Pacific | | | | | | |
| 1 Least deprived | 109.2 (104.8, 113.5) | Ref. | | 44.7 (35.6, 53.9) | Ref. | |
| 2 | 107.5 (103.5, 111.5) | 0.99 (0.93, 1.04) | 0.59 | 37.7 (34.0, 41.4) | 0.84 (0.67, 1.06) | 0.14 |
| 3 | 111.6 (107.4, 115.8) | 1.02 (0.97, 1.08) | 0.42 | 42.6 (36.5, 48.8) | 0.95 (0.74, 1.22) | 0.70 |
| 4 | 116.8 (113.1, 120.5) | 1.07 (1.02, 1.13) | <0.01 | 45.6 (41.4, 49.8) | 1.02 (0.82, 1.27) | 0.87 |
| 5 Most deprived | 133.2 (129.1, 137.3) | 1.22 (1.16, 1.28) | <0.01 | 45.8 (41.5, 50.1) | 1.02 (0.82, 1.28) | 0.84 |
| Europe (excluding UK) & Central Asia | | | | | | |
| 1 Least deprived | 109.5 (106.7, 112.3) | Ref. | | 43.9 (37.6, 50.2) | Ref. | |
| 2 | 109.4 (106.9, 111.9) | 1.00 (0.97, 1.03) | 0.96 | 42.2 (39.3, 45.0) | 0.96 (0.82, 1.12) | 0.61 |
| 3 | 108.7 (106.6, 110.8) | 0.99 (0.96, 1.02) | 0.65 | 41.7 (39.6, 43.9) | 0.95 (0.82, 1.10) | 0.50 |
| 4 | 111.1 (109.3, 112.8) | 1.01 (0.99, 1.04) | 0.34 | 40.9 (39.1, 42.7) | 0.93 (0.81, 1.08) | 0.33 |
| 5 Most deprived | 127.8 (125.8, 129.8) | 1.17 (1.13, 1.20) | <0.01 | 44.9 (43.0, 46.7) | 1.02 (0.89, 1.18) | 0.77 |
| Latin America & Caribbean | | | | | | |
| 1 Least deprived | 119.3 (110.4, 128.2) | Ref. | | 51.2 (38.8, 63.6) | Ref. | |
| 2 | 114.1 (105.5, 122.8) | 0.96 (0.86, 1.06) | 0.41 | 51.1 (34.3, 68.0) | 1.00 (0.66, 1.50) | 0.99 |
| 3 | 116.4 (109.9, 123.0) | 0.98 (0.89, 1.07) | 0.61 | 48.2 (40.6, 55.9) | 0.94 (0.71, 1.26) | 0.68 |
| 4 | 111.2 (106.0, 116.3) | 0.93 (0.85, 1.02) | 0.11 | 49.6 (44.4, 54.8) | 0.97 (0.74, 1.26) | 0.81 |
| 5 Most deprived | 123.8 (118.8, 128.9) | 1.04 (0.95, 1.13) | 0.39 | 51.6 (47.2, 56.0) | 1.01 (0.78, 1.30) | 0.96 |
| Middle East & North Africa | | | | | | |
| 1 Least deprived | 116.4 (107.7, 125.1) | Ref. | | 40.3 (34.3, 46.3) | Ref. | |
| 2 | 107.5 (100.6, 114.4) | 0.92 (0.84, 1.02) | 0.11 | 49.2 (39.7, 58.8) | 1.22 (0.96, 1.56) | 0.10 |
| 3 | 108.9 (103.2, 114.6) | 0.94 (0.85, 1.02) | 0.15 | 48.9 (43.7, 54.1) | 1.21 (1.01, 1.45) | 0.04 |
| 4 | 124.5 (119.5, 129.5) | 1.07 (0.98, 1.16) | 0.12 | 60.0 (45.9, 74.1) | 1.49 (1.13, 1.96) | <0.01 |
| 5 Most deprived | 150.9 (146.5, 155.3) | 1.30 (1.20, 1.40) | <0.01 | 63.9 (56.9, 70.8) | 1.59 (1.32, 1.90) | <0.01 |
| North America | | | | | | |
| 1 Least deprived | 109.9 (102.5, 117.4) | Ref. | | 46.3 (36.4, 56.1) | Ref. | |
| 2 | 89.8 (83.8, 95.8) | 0.82 (0.74, 0.90) | <0.01 | 32.3 (27.9, 36.6) | 0.70 (0.54, 0.90) | <0.01 |

| | | | | | | |
|---|---|---|---|---|---|---|
| 3 | 100.9 (92.6, 109.2) | 0.92 (0.83, 1.02) | 0.11 | 37.2 (31.0, 43.5) | 0.80 (0.61, 1.05) | 0.11 |
| 4 | 98.4 (91.0, 105.9) | 0.90 (0.81, 0.99) | 0.03 | 39.1 (32.9, 45.3) | 0.84 (0.65, 1.10) | 0.21 |
| 5 Most deprived | 128.2 (114.5, 141.8) | 1.17 (1.03, 1.32) | 0.02 | 56.6 (37.2, 76.0) | 1.22 (0.82, 1.83) | 0.33 |
| South Asia | | | | | | |
| 1 Least deprived | 135.7 (130.7, 140.7) | Ref. | | 46.6 (41.7, 51.4) | Ref. | |
| 2 | 139.1 (134.6, 143.5) | 1.02 (0.98, 1.08) | 0.32 | 47.4 (43.5, 51.3) | 1.02 (0.89, 1.16) | 0.79 |
| 3 | 140.1 (136.6, 143.6) | 1.03 (0.99, 1.08) | 0.16 | 51.9 (48.7, 55.1) | 1.11 (0.99, 1.25) | 0.07 |
| 4 | 145.7 (143.0, 148.4) | 1.07 (1.03, 1.12) | <0.01 | 52.7 (49.4, 56.1) | 1.13 (1.00, 1.28) | 0.04 |
| 5 Most deprived | 180.7 (178.2, 183.2) | 1.33 (1.28, 1.38) | <0.01 | 64.4 (60.5, 68.4) | 1.38 (1.23, 1.56) | <0.01 |
| Sub-Saharan Africa | | | | | | |
| 1 Least deprived | 115.6 (111.2, 120.0) | Ref. | | 43.8 (39.0, 48.5) | Ref. | |
| 2 | 118.2 (113.9, 122.6) | 1.02 (0.97, 1.08) | 0.40 | 46.1 (42.0, 50.3) | 1.05 (0.92, 1.21) | 0.46 |
| 3 | 117.1 (113.5, 120.8) | 1.01 (0.97, 1.06) | 0.59 | 44.8 (41.6, 48.1) | 1.02 (0.90, 1.16) | 0.72 |
| 4 | 114.0 (111.5, 116.5) | 0.99 (0.94, 1.03) | 0.53 | 49.9 (46.6, 53.2) | 1.14 (1.01, 1.29) | 0.04 |
| 5 Most deprived | 127.2 (125.1, 129.4) | 1.10 (1.06, 1.15) | <0.01 | 52.1 (49.9, 54.3) | 1.19 (1.06, 1.33) | <0.01 |
| UK | | | | | | |
| 1 Least deprived | 147.3 (146.1, 148.5) | Ref. | | 46.4 (45.2, 47.7) | Ref. | |
| 2 | 159.5 (158.3, 160.8) | 1.08 (1.07, 1.09) | <0.01 | 47.6 (46.3, 48.9) | 1.02 (1.00, 1.05) | 0.09 |
| 3 | 168.0 (166.8, 169.3) | 1.14 (1.13, 1.15) | <0.01 | 48.6 (47.3, 49.9) | 1.05 (1.02, 1.08) | <0.01 |
| 4 | 183.4 (182.1, 184.7) | 1.25 (1.23, 1.26) | <0.01 | 50.6 (49.4, 51.8) | 1.09 (1.06, 1.12) | <0.01 |
| 5 Most deprived | 210.8 (209.3, 212.2) | 1.43 (1.42, 1.44) | <0.01 | 54.6 (53.3, 55.8) | 1.18 (1.14, 1.21) | <0.01 |

CI = Confidence interval, IMD = index of multiple deprivation, IRR = incidence rate ratio; *results derived from negative binomial regression models adjusted for year of birth, maternal region of birth, IMD group and maternal region of birth*IMD group interaction terms (regression model results available on request); marginal incidence rates derived from models with year of birth set to mid-study (2011); IRR of admission rates for IMD groups in comparison to the least deprived IMD group, within maternal region groups; ***N* = 4,174,596, AIC = 7796925.19 (compared with AIC = 7797823.04 for model without interaction term); ****N* = 4,174,596, AIC = 3291383.78 (compared with AIC = 3291546.74 for model without interaction term)

**Supplementary Table 8. Estimated incidence rates and IRRs of emergency and planned hospital admissions, by maternal country of birth and IMD group (derived from negative binomial regression models)***

| | Emergency admissions** Incidence rate (95% CI) per 1000 child-years | IRR (95% CI) | p-value | Planned admissions*** Incidence rate (95% CI) per 1000 child-years | IRR (95% CI) | p-value |
|---|---|---|---|---|---|---|
| Bangladesh | | | | | | |
| 1 Least deprived | 135.6 (119.3, 151.9) | Ref. | | 43.1 (29.2, 57.0) | Ref. | |
| 2 | 143.4 (128.0, 158.8) | 1.06 (0.90, 1.24) | 0.49 | 53.4 (41.1, 65.6) | 1.24 (0.83, 1.84) | 0.29 |
| 3 | 153.9 (142.9, 164.9) | 1.14 (0.99, 1.30) | 0.07 | 55.6 (47.2, 64.1) | 1.29 (0.90, 1.84) | 0.16 |
| 4 | 146.0 (137.6, 154.4) | 1.08 (0.94, 1.23) | 0.28 | 50.0 (44.9, 55.0) | 1.16 (0.83, 1.62) | 0.39 |
| 5 Most deprived | 158.2 (154.1, 162.3) | 1.17 (1.03, 1.32) | 0.01 | 60.4 (55.9, 64.9) | 1.40 (1.01, 1.95) | 0.05 |
| India | | | | | | |
| 1 Least deprived | 132.4 (125.9, 138.9) | Ref. | | 40.9 (36.2, 45.7) | Ref. | |
| 2 | 130.5 (124.5, 136.5) | 0.99 (0.92, 1.05) | 0.67 | 39.0 (34.3, 43.6) | 0.95 (0.81, 1.12) | 0.56 |
| 3 | 124.8 (120.5, 129.1) | 0.94 (0.89, 1.00) | 0.05 | 41.3 (37.5, 45.0) | 1.01 (0.87, 1.17) | 0.92 |
| 4 | 121.9 (118.4, 125.5) | 0.92 (0.87, 0.97) | <0.01 | 41.3 (37.9, 44.8) | 1.01 (0.88, 1.16) | 0.90 |
| 5 Most deprived | 145.5 (141.0, 150.1) | 1.10 (1.04, 1.16) | <0.01 | 42.3 (38.5, 46.2) | 1.03 (0.89, 1.20) | 0.66 |
| Nigeria | | | | | | |
| 1 Least deprived | 114.2 (100.0, 128.3) | Ref. | | 42.0 (33.6, 50.4) | Ref. | |
| 2 | 111.3 (101.4, 121.3) | 0.98 (0.84, 1.14) | 0.75 | 44.2 (35.4, 52.9) | 1.05 (0.80, 1.39) | 0.73 |
| 3 | 106.8 (98.7, 114.9) | 0.94 (0.81, 1.08) | 0.37 | 41.8 (34.8, 48.8) | 1.00 (0.77, 1.29) | 0.97 |
| 4 | 107.6 (102.9, 112.3) | 0.94 (0.83, 1.07) | 0.38 | 53.3 (47.9, 58.6) | 1.27 (1.02, 1.58) | 0.04 |
| 5 Most deprived | 114.5 (110.5, 118.6) | 1.00 (0.88, 1.14) | 0.96 | 48.7 (43.6, 53.8) | 1.16 (0.93, 1.45) | 0.19 |
| Pakistan | | | | | | |
| 1 Least deprived | 154.7 (142.7, 166.7) | Ref. | | 61.3 (50.0, 72.6) | Ref. | |
| 2 | 161.3 (151.6, 171.1) | 1.04 (0.95, 1.15) | 0.40 | 58.9 (49.0, 68.8) | 0.96 (0.75, 1.23) | 0.75 |
| 3 | 164.6 (156.6, 172.7) | 1.06 (0.97, 1.17) | 0.18 | 65.8 (57.9, 73.7) | 1.07 (0.86, 1.34) | 0.52 |
| 4 | 176.5 (171.2, 181.9) | 1.14 (1.05, 1.24) | <0.01 | 61.5 (53.8, 69.2) | 1.00 (0.80, 1.25) | 0.97 |
| 5 Most deprived | 209.8 (205.7, 213.9) | 1.36 (1.25, 1.47) | <0.01 | 75.1 (67.8, 82.3) | 1.22 (1.00, 1.51) | 0.06 |
| Poland | | | | | | |
| 1 Least deprived | 98.3 (93.1, 103.4) | Ref. | | 38.9 (34.6, 43.2) | Ref. | |
| 2 | 100.0 (95.7, 104.3) | 1.02 (0.95, 1.09) | 0.60 | 40.3 (35.2, 45.4) | 1.04 (0.88, 1.22) | 0.68 |

| | | | | | | |
|---|---|---|---|---|---|---|
| 3 | 99.9 (96.5, 103.4) | 1.02 (0.96, 1.08) | 0.59 | 40.1 (36.4, 43.8) | 1.03 (0.89, 1.19) | 0.68 |
| 4 | 102.6 (99.9, 105.2) | 1.04 (0.99, 1.11) | 0.15 | 39.6 (36.3, 42.9) | 1.02 (0.89, 1.17) | 0.80 |
| 5 Most deprived | 116.4 (113.4, 119.4) | 1.18 (1.12, 1.25) | <0.01 | 40.1 (37.5, 42.7) | 1.03 (0.91, 1.17) | 0.63 |
| UK | | | | | | |
| 1 Least deprived | 147.6 (146.4, 148.8) | Ref. | | 46.4 (45.1, 47.6) | Ref. | |
| 2 | 159.8 (158.5, 161.1) | 1.08 (1.07, 1.09) | <0.01 | 47.6 (46.2, 48.9) | 1.03 (1.00, 1.06) | 0.09 |
| 3 | 168.3 (167.0, 169.6) | 1.14 (1.13, 1.15) | <0.01 | 48.6 (47.3, 49.9) | 1.05 (1.02, 1.08) | <0.01 |
| 4 | 183.7 (182.3, 185.1) | 1.24 (1.23, 1.26) | <0.01 | 50.5 (49.3, 51.8) | 1.09 (1.06, 1.12) | <0.01 |
| 5 Most deprived | 211.1 (209.6, 212.7) | 1.43 (1.42, 1.44) | <0.01 | 54.5 (53.2, 55.8) | 1.18 (1.14, 1.21) | <0.01 |

CI = Confidence interval, IMD = index of multiple deprivation, IRR = incidence rate ratio; *results derived from negative binomial regression models adjusted for year of birth, maternal country of birth, IMD group and maternal country of birth*IMD group interaction terms (regression model results available on request); marginal incidence rates derived from models with year of birth set to mid-study (2011); IRR of admission rates for IMD groups in comparison to the least deprived IMD group, within maternal country groups; ***N* = 3,492,139, AIC = 6729769.86 (compared with AIC = 6730123.41 for model without interaction term); ****N* = 3,492,139, AIC = 2775796.38 (compared with AIC = 2775829.28 for model without interaction term)

**Supplementary Table 9. Estimated incidence rates and IRRs of emergency and planned hospital admissions, by parental migration status and IMD group (derived from negative binomial regression models*)**

| | Emergency admissions** Incidence rate (95% CI) per 1000 child-years | IRR (95% CI) | *p*-value | Planned admissions*** Incidence rate (95% CI) per 1000 child-years | IRR (95% CI) | *p*-value |
|---|---|---|---|---|---|---|
| Mother UK-born & SP UK-born | | | | | | |
| 1 Least deprived | 146.9 (145.7, 148.1) | Ref. | | 46.4 (45.1, 47.7) | Ref. | |
| 2 | 158.5 (157.3, 159.8) | 1.08 (1.07, 1.09) | <0.01 | 47.5 (46.2, 48.9) | 1.03 (0.99, 1.06) | 0.12 |
| 3 | 166.4 (165.1, 167.7) | 1.13 (1.12, 1.14) | <0.01 | 48.2 (46.8, 49.5) | 1.04 (1.01, 1.07) | 0.01 |
| 4 | 182.0 (180.7, 183.4) | 1.24 (1.23, 1.25) | <0.01 | 49.7 (48.4, 51.0) | 1.07 (1.04, 1.10) | <0.01 |
| 5 Most deprived | 208.3 (206.8, 209.8) | 1.42 (1.41, 1.43) | <0.01 | 53.0 (51.7, 54.4) | 1.14 (1.11, 1.18) | <0.01 |
| Mother UK-born & SP non-UK-born | | | | | | |
| 1 Least deprived | 130.4 (126.9, 134.0) | Ref. | | 45.6 (42.3, 48.9) | Ref. | |
| 2 | 140.7 (137.0, 144.4) | 1.08 (1.04, 1.12) | <0.01 | 44.7 (42.0, 47.5) | 0.98 (0.89, 1.08) | 0.68 |
| 3 | 150.1 (146.5, 153.7) | 1.15 (1.11, 1.19) | <0.01 | 50.1 (46.8, 53.3) | 1.10 (1.00, 1.21) | 0.05 |
| 4 | 163.0 (159.3, 166.7) | 1.25 (1.21, 1.29) | <0.01 | 52.6 (49.6, 55.6) | 1.15 (1.06, 1.26) | <0.01 |
| 5 Most deprived | 199.5 (195.9, 203.0) | 1.53 (1.48, 1.58) | <0.01 | 62.1 (58.9, 65.3) | 1.36 (1.25, 1.48) | <0.01 |
| Mother UK-born (no SP registered) | | | | | | |
| 1 Least deprived | 209.6 (201.8, 217.5) | Ref. | | 51.1 (45.8, 56.4) | Ref. | |
| 2 | 221.5 (214.9, 228.1) | 1.06 (1.01, 1.11) | 0.02 | 54.2 (49.8, 58.7) | 1.06 (0.93, 1.21) | 0.37 |
| 3 | 223.0 (217.3, 228.6) | 1.06 (1.02, 1.11) | 0.01 | 55.0 (50.9, 59.2) | 1.08 (0.95, 1.22) | 0.25 |
| 4 | 223.7 (219.4, 228.0) | 1.07 (1.02, 1.11) | <0.01 | 58.4 (55.3, 61.5) | 1.14 (1.02, 1.28) | 0.02 |
| 5 Most deprived | 239.4 (235.9, 242.8) | 1.14 (1.10, 1.19) | <0.01 | 58.3 (55.1, 61.4) | 1.14 (1.02, 1.28) | 0.02 |
| Mother non-UK-born & SP non-UK-born | | | | | | |
| 1 Least deprived | 109.9 (107.5, 112.3) | Ref. | | 41.1 (38.7, 43.4) | Ref. | |
| 2 | 108.9 (106.9, 111.0) | 0.99 (0.96, 1.02) | 0.54 | 42.6 (40.2, 45.0) | 1.04 (0.96, 1.12) | 0.35 |
| 3 | 113.0 (111.3, 114.7) | 1.03 (1.00, 1.06) | 0.03 | 44.4 (42.7, 46.1) | 1.08 (1.01, 1.15) | 0.02 |
| 4 | 118.2 (116.7, 119.6) | 1.08 (1.05, 1.10) | <0.01 | 47.4 (45.5, 49.3) | 1.15 (1.08, 1.23) | <0.01 |
| 5 Most deprived | 141.3 (139.9, 142.7) | 1.29 (1.26, 1.32) | <0.01 | 54.0 (52.1, 56.0) | 1.32 (1.23, 1.40) | <0.01 |
| Mother non-UK-born & SP UK-born | | | | | | |
| 1 Least deprived | 120.8 (118.0, 123.6) | Ref. | | 49.1 (42.7, 55.5) | Ref. | |
| 2 | 124.4 (121.5, 127.3) | 1.03 (1.00, 1.06) | 0.07 | 44.6 (41.6, 47.5) | 0.91 (0.79, 1.04) | 0.18 |

| | | | | | | |
|---|---|---|---|---|---|---|
| 3 | 128.8 (125.9, 131.8) | 1.07 (1.03, 1.10) | <0.01 | 47.2 (43.6, 50.7) | 0.96 (0.83, 1.11) | 0.59 |
| 4 | 138.6 (135.8, 141.5) | 1.15 (1.11, 1.18) | <0.01 | 47.7 (43.6, 51.8) | 0.97 (0.84, 1.13) | 0.72 |
| 5 Most deprived | 174.5 (171.0, 177.9) | 1.44 (1.40, 1.49) | <0.01 | 59.4 (55.3, 63.4) | 1.21 (1.05, 1.39) | 0.01 |
| Mother non-UK-born (sole registration) | | | | | | |
| 1 Least deprived | 120.0 (105.4, 134.7) | Ref. | | 28.6 (20.9, 36.4) | Ref. | |
| 2 | 132.5 (120.9, 144.1) | 1.10 (0.95, 1.28) | 0.20 | 44.2 (35.1, 53.3) | 1.54 (1.10, 2.16) | 0.01 |
| 3 | 134.6 (126.3, 143.0) | 1.12 (0.98, 1.29) | 0.10 | 46.9 (40.1, 53.6) | 1.64 (1.21, 2.22) | <0.01 |
| 4 | 124.4 (119.3, 129.5) | 1.04 (0.91, 1.18) | 0.59 | 51.9 (46.3, 57.4) | 1.81 (1.36, 2.42) | <0.01 |
| 5 Most deprived | 141.2 (136.7, 145.8) | 1.18 (1.04, 1.33) | 0.01 | 50.2 (46.4, 54.0) | 1.75 (1.33, 2.32) | <0.01 |

CI = Confidence interval, IMD = index of multiple deprivation, IRR = incidence rate ratio, SP = second parent *results derived from negative binomial regression models adjusted for year of birth, maternal region of birth, IMD group and maternal region of birth*IMD group interaction terms (regression model results available on request); marginal incidence rates derived from models with year of birth set to mid-study (2011); IRR of admission rates for IMD groups in comparison to the least deprived IMD group, within maternal region groups; ***N* = 4,174,596, AIC = 7796685.71 (compared with AIC = 7797359.99 for model without interaction term); ****N* = 4,174,596, AIC = 3291959.24 (compared with AIC = 3292159.78 for model without interaction term)

#### 6. Appendix F. Results: Secondary outcomes

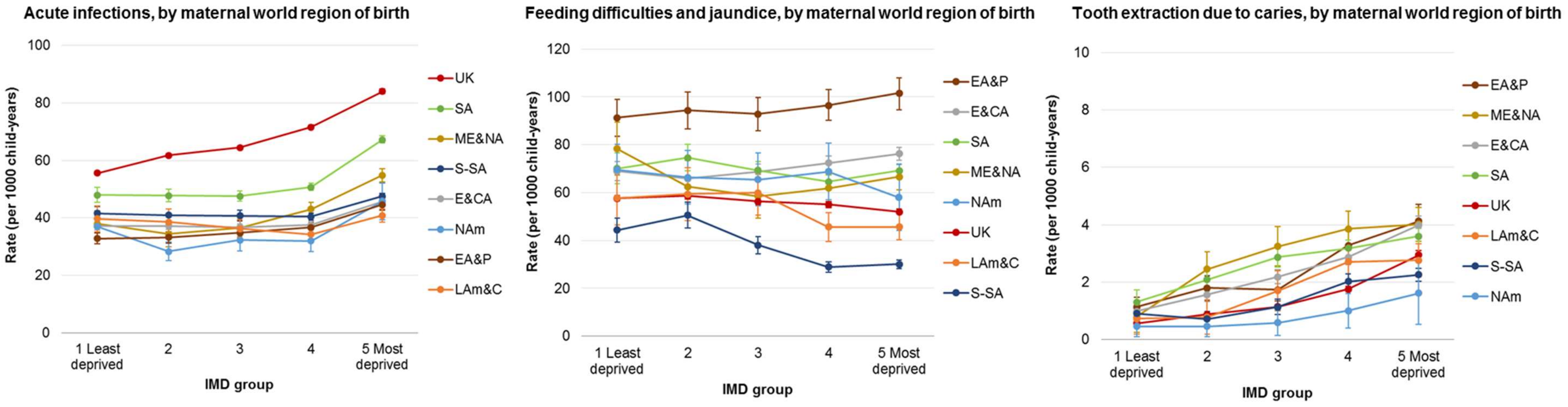


**Supplementary Figure 2. Estimated incidence rates of admissions for acute infections, feed difficulties and jaundice and tooth extractions for caries per 1000 person-years, by maternal world region of birth and IMD group (see Supplementary Table 10 for further details)**; IMD= index of multiple deprivation; EA&P= East Asia and Pacific, E&CA= Europe (excl. UK) and Central Asia, LAm&C= Latin America and Caribbean, ME&NA= Middle East and North Africa, NAm= North America, SA= South Asia, S-SA= Sub-Saharan Africa. **Note. Y-axis scales are different**

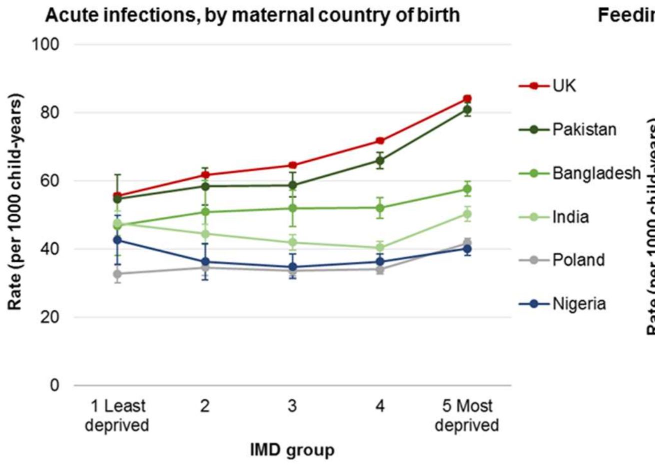
Acute infections, by maternal country of birth
Rate (per 1000 child-years)
100
80
60
40
20
0
1 Least deprived
2
3
4
5 Most deprived
IMD group
UK
Pakistan
Bangladesh
India
Poland
Nigeria

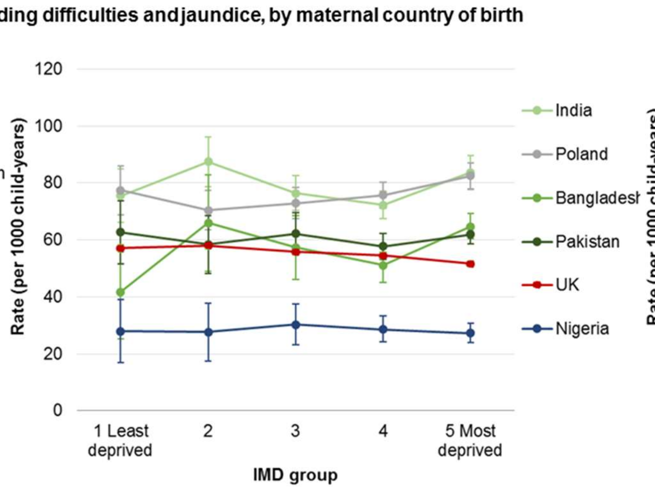
Feeding difficulties and jaundice, by maternal country of birth
Rate (per 1000 child-years)
120
100
80
60
40
20
0
1 Least deprived
2
3
4
5 Most deprived
IMD group
India
Poland
Bangladesh
Pakistan
UK
Nigeria

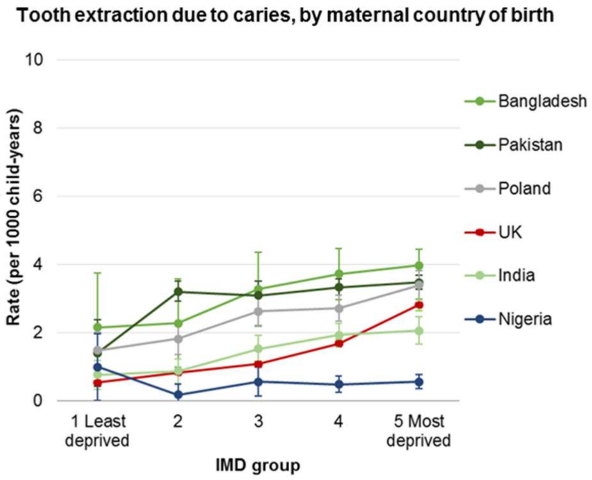
Tooth extraction due to caries, by maternal country of birth
Rate (per 1000 child-years)
10
8
6
4
2
0
1 Least deprived
2
3
4
5 Most deprived
IMD group
Bangladesh
Pakistan
Poland
UK
India
Nigeria

**Supplementary Figure 3.** Estimated incidence rates of emergency and planned hospital admissions per 1000 person-years, by maternal country of birth/parental migration status and IMD group (see Supplementary Table 11 for further details); IMD= index of multiple deprivation. **Note. Y-axis scales are different**

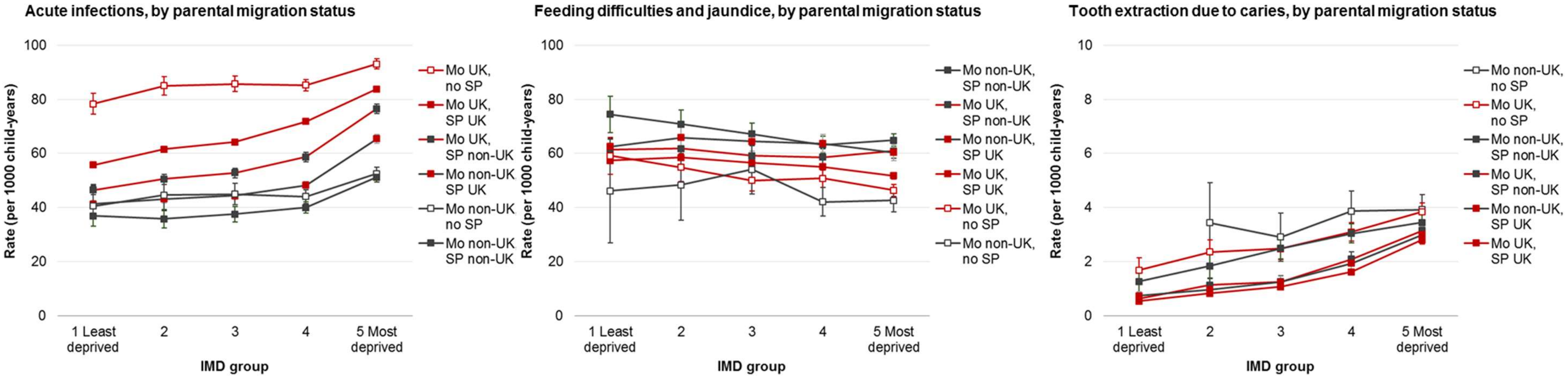


**Supplementary Figure 4.** Estimated incidence rates of emergency and planned hospital admissions per 1000 person-years, by maternal country of birth/parental migration status and IMD group (see Supplementary Table 12 for further details); IMD= index of multiple deprivation; Mo = mother, SP = second parent, UK = UK-born parent, Non-UK=non-UK-born parent. **Note. Y-axis scales are different**

**Supplementary Table 10. Estimated incidence rates (per 1000 child-years) and IRRs of hospital admissions with specified diagnoses, by maternal region of birth and IMD group (derived from negative binomial/Poisson regression models)***

| | Acute infection** | | | Neonatal feeding difficulties*** | | | Tooth extraction for caries[a] | | |
|---|---|---|---|---|---|---|---|---|---|
| | Incidence rate (95% CI) | IRR (95% CI) | p-value | Incidence rate (95% CI) | IRR (95% CI) | p-value | Incidence rate (95% CI) | IRR (95% CI) | p-value |
| East-Asia & Pacific | | | | | | | | | |
| 1 Least deprived | 32.8 (30.9,34.8) | Ref. | | 91.3 (83.6,98.9) | Ref. | | 1.13 (0.78, 1.48) | Ref. | |
| 2 | 33.2 (31.2,35.1) | 1.01 (0.93,1.10) | 0.82 | 94.4 (86.5,102.3) | 1.03 (0.92,1.16) | 0.57 | 1.80 (1.36, 2.23) | 1.59 (1.08,2.34) | 0.02 |
| 3 | 34.8 (32.8,36.7) | 1.06 (0.97,1.15) | 0.18 | 92.8 (85.9,99.7) | 1.02 (0.91,1.14) | 0.76 | 1.73 (1.35, 2.12) | 1.53 (1.05,2.23) | 0.03 |
| 4 | 36.7 (35.0,38.5) | 1.12 (1.04,1.21) | <0.01 | 96.5 (90.1,103.0) | 1.06 (0.95,1.17) | 0.30 | 3.28 (2.77, 3.79) | 2.90 (2.07,4.07) | <0.01 |
| 5 Most deprived | 44.6 (42.6,46.6) | 1.36 (1.26,1.46) | <0.01 | 101.6 (94.5,108.7) | 1.11 (1.00,1.24) | 0.05 | 4.13 (3.55, 4.72) | 3.66 (2.62,5.10) | <0.01 |
| Europe (excl. UK) & Central Asia | | | | | | | | | |
| 1 Least deprived | 37.1 (35.7,38.4) | Ref. | | 68.9 (64.9,72.9) | Ref. | | 0.98 (0.78, 1.18) | Ref. | |
| 2 | 37.2 (36.0,38.5) | 1.00 (0.96,1.05) | 0.85 | 65.8 (62.3,69.2) | 0.95 (0.88,1.03) | 0.22 | 1.56 (1.32, 1.81) | 1.60 (1.24,2.05) | <0.01 |
| 3 | 36.8 (35.8,37.9) | 0.99 (0.95,1.04) | 0.79 | 68.8 (65.7,71.9) | 1.00 (0.93,1.07) | 0.97 | 2.19 (1.94, 2.44) | 2.23 (1.78,2.81) | <0.01 |
| 4 | 37.5 (36.6,38.4) | 1.01 (0.97,1.06) | 0.62 | 72.4 (69.6,75.2) | 1.05 (0.98,1.12) | 0.14 | 2.87 (2.61, 3.13) | 2.93 (2.36,3.64) | <0.01 |
| 5 Most deprived | 45.8 (44.8,46.9) | 1.24 (1.19,1.29) | <0.01 | 76.2 (73.5,79.0) | 1.11 (1.04,1.18) | <0.01 | 3.99 (3.67, 4.31) | 4.08 (3.30,5.04) | <0.01 |
| Latin America & Caribbean | | | | | | | | | |
| 1 Least deprived | 39.7 (35.2,44.2) | Ref. | | 57.7 (46.6,68.9) | Ref. | | 0.73 (0.19, 1.27) | Ref. | |
| 2 | 38.6 (34.2,43.1) | 0.97 (0.83,1.14) | 0.74 | 59.4 (48.4,70.4) | 1.03 (0.79,1.34) | 0.84 | 0.78 (0.18, 1.38) | 1.07 (0.37,3.11) | 0.91 |
| 3 | 36.3 (33.2,39.4) | 0.91 (0.79,1.05) | 0.21 | 59.9 (50.7,69.1) | 1.04 (0.81,1.33) | 0.77 | 1.70 (1.01, 2.39) | 2.33 (1.01,5.41) | 0.05 |
| 4 | 34.3 (31.9,36.6) | 0.86 (0.76,0.99) | 0.03 | 45.6 (39.6,51.7) | 0.79 (0.63,1.00) | 0.05 | 2.71 (2.10, 3.32) | 3.72 (1.72,8.05) | <0.01 |
| 5 Most deprived | 40.8 (38.4,43.1) | 1.03 (0.90,1.17) | 0.68 | 45.6 (40.2,51.0) | 0.79 (0.63,0.99) | 0.04 | 2.76 (2.17, 3.34) | 3.79 (1.76,8.16) | <0.01 |
| Middle East & North Africa | | | | | | | | | |
| 1 Least deprived | 37.9 (33.7,42.2) | Ref. | | 78.3 (65.1,91.6) | Ref. | | 0.79 (0.18, 1.40) | Ref. | |
| 2 | 34.4 (31.1,37.7) | 0.91 (0.78,1.05) | 0.19 | 62.5 (52.7,72.3) | 0.80 (0.63,1.00) | 0.05 | 2.45 (1.62, 3.28) | 3.11 (1.34,7.23) | 0.01 |
| 3 | 36.5 (33.8,39.2) | 0.96 (0.84,1.10) | 0.57 | 58.5 (50.9,66.2) | 0.75 (0.60,0.92) | 0.01 | 3.25 (2.47, 4.02) | 4.12 (1.84,9.25) | <0.01 |
| 4 | 43.0 (40.6,45.4) | 1.13 (1.00,1.28) | 0.05 | 61.8 (55.6,68.1) | 0.79 (0.65,0.96) | 0.02 | 3.86 (3.19, 4.52) | 4.89 (2.22,10.79) | <0.01 |
| 5 Most deprived | 54.9 (52.7,57.2) | 1.45 (1.29,1.63) | <0.01 | 66.6 (61.6,71.6) | 0.85 (0.71,1.02) | 0.08 | 4.02 (3.49, 4.55) | 5.10 (2.33,11.16) | <0.01 |
| North America | | | | | | | | | |
| 1 Least deprived | 37.0 (33.2,40.8) | Ref. | | 69.6 (58.9,80.3) | Ref. | | 0.46 (0.09, 0.82) | Ref. | |
| 2 | 28.3 (25.2,31.5) | 0.77 (0.66,0.89) | <0.01 | 66.4 (55.2,77.7) | 0.95 (0.76,1.20) | 0.69 | 0.46 (0.09, 0.83) | 1.00 (0.32,3.11) | 0.99 |
| 3 | 32.3 (28.4,36.2) | 0.87 (0.75,1.02) | 0.09 | 65.4 (54.4,76.5) | 0.94 (0.75,1.18) | 0.59 | 0.59 (0.15, 1.02) | 1.28 (0.43,3.80) | 0.66 |

| | | | | | | | | | |
|---|---|---|---|---|---|---|---|---|---|
| 4 | 32.0 (28.2,35.8) | 0.86 (0.74,1.01) | 0.07 | 68.8 (57.0,80.6) | 0.99 (0.79,1.24) | 0.92 | 1.01 (0.41, 1.61) | 2.21 (0.82,5.98) | 0.12 |
| 5 Most deprived | 45.7 (39.3,52.1) | 1.23 (1.04,1.47) | 0.02 | 58.0 (44.1,71.9) | 0.83 (0.63,1.11) | 0.21 | 1.62 (0.52, 2.72) | 3.55 (1.25,10.10) | 0.02 |
| South Asia | | | | | | | | | |
| 1 Least deprived | 48.0 (45.4,50.6) | Ref. | | 70.1 (63.7,76.5) | Ref. | | 1.31 (0.90, 1.72) | Ref. | |
| 2 | 47.7 (45.5,49.9) | 0.99 (0.92,1.07) | 0.84 | 74.6 (69.1,80.1) | 1.06 (0.95,1.19) | 0.29 | 2.09 (1.68, 2.50) | 1.60 (1.11,2.30) | 0.01 |
| 3 | 47.5 (45.8,49.3) | 0.99 (0.93,1.06) | 0.75 | 69.3 (65.5,73.1) | 0.99 (0.89,1.10) | 0.83 | 2.87 (2.52, 3.23) | 2.20 (1.58,3.06) | <0.01 |
| 4 | 50.7 (49.5,52.0) | 1.06 (1.00,1.12) | 0.07 | 64.5 (61.7,67.2) | 0.92 (0.83,1.02) | 0.10 | 3.18 (2.89, 3.47) | 2.43 (1.77,3.35) | <0.01 |
| 5 Most deprived | 67.2 (66.0,68.5) | 1.40 (1.32,1.48) | <0.01 | 69.2 (66.8,71.6) | 0.99 (0.90,1.09) | 0.80 | 3.60 (3.33, 3.87) | 2.75 (2.01,3.77) | <0.01 |
| Sub-Saharan Africa | | | | | | | | | |
| 1 Least deprived | 41.6 (39.4,43.9) | Ref. | | 44.4 (39.4,49.4) | Ref. | | 0.91 (0.62, 1.21) | Ref. | |
| 2 | 41.0 (38.8,43.2) | 0.98 (0.91,1.06) | 0.69 | 50.6 (45.2,56.0) | 1.14 (0.98,1.33) | 0.09 | 0.72 (0.46, 0.98) | 0.79 (0.49,1.26) | 0.32 |
| 3 | 40.7 (38.9,42.6) | 0.98 (0.91,1.05) | 0.53 | 38.0 (34.4,41.6) | 0.86 (0.74,0.99) | 0.04 | 1.14 (0.88, 1.40) | 1.25 (0.84,1.84) | 0.27 |
| 4 | 40.4 (39.1,41.6) | 0.97 (0.91,1.03) | 0.33 | 28.9 (26.7,31.1) | 0.65 (0.57,0.74) | <0.01 | 2.02 (1.76, 2.28) | 2.21 (1.58,3.11) | <0.01 |
| 5 Most deprived | 47.5 (46.4,48.6) | 1.14 (1.08,1.21) | <0.01 | 30.1 (28.3,31.9) | 0.68 (0.60,0.77) | <0.01 | 2.26 (2.04, 2.49) | 2.47 (1.78,3.44) | <0.01 |
| UK | | | | | | | | | |
| 1 Least deprived | 55.6 (55.0,56.2) | Ref. | | 57.6 (56.4,58.9) | Ref. | | 0.57 (0.52, 0.62) | Ref. | |
| 2 | 61.7 (61.1,62.4) | 1.11 (1.10,1.12) | <0.01 | 58.6 (57.3,59.8) | 1.02 (0.99,1.04) | 0.19 | 0.88 (0.82, 0.95) | 1.56 (1.42,1.71) | <0.01 |
| 3 | 64.5 (63.8,65.1) | 1.16 (1.15,1.17) | <0.01 | 56.3 (55.1,57.6) | 0.98 (0.95,1.00) | 0.06 | 1.15 (1.07, 1.22) | 2.02 (1.85,2.20) | <0.01 |
| 4 | 71.6 (70.9,72.3) | 1.29 (1.27,1.30) | <0.01 | 55.0 (53.8,56.2) | 0.95 (0.93,0.98) | <0.01 | 1.76 (1.66, 1.86) | 3.10 (2.86,3.36) | <0.01 |
| 5 Most deprived | 84.0 (83.3,84.8) | 1.51 (1.49,1.53) | <0.01 | 52.0 (51.0,53.1) | 0.90 (0.88,0.92) | <0.01 | 2.95 (2.80, 3.11) | 5.21 (4.82,5.62) | <0.01 |

CI = Confidence interval, IMD = index of multiple deprivation, IRR = incidence rate ratio; *results derived from negative binomial/Poisson regression models adjusted for year of birth, maternal region of birth, IMD group and maternal region of birth*IMD group interaction term (regression model results available on request); marginal incidence rates derived from models with year of birth set to mid-study (2011); IRR of admission rates for IMD groups in comparison to the least deprived IMD group, within maternal region groups; **Negative binomial regression, *N* = 4,174,596, AIC = 4206765.23 (compared with AIC = 4207230.40 for model without interaction term); *** Negative binomial regression, *N* = 4,174,596, AIC = 1088294.95 (compared with AIC = 1088471.92 for model without interaction term); [a]Poisson regression, *N* = 2,973,284, AIC = 213907.46 (compared with AIC = 214088.94 for model without interaction term)

**Supplementary Table 11. Estimated incidence rates (per 1000 child-years) and IRRs of hospital admissions with specified diagnoses, by maternal country of birth and IMD group (derived from negative binomial/Poisson regression models)***

| | Acute infections** | | | Feeding difficulties and jaundice*** | | | Tooth extractions for caries[a] | | |
|---|---|---|---|---|---|---|---|---|---|
| | Incidence rate (95% CI) | IRR (95% CI) | p-value | Incidence rate (95% CI) | IRR (95% CI) | p-value | Incidence rate (95% CI) | IRR (95% CI) | p-value |
| Bangladesh | | | | | | | | | |
| 1 Least deprived | 46.9 (38.2,55.6) | Ref. | | 41.7 (25.3,58.2) | Ref. | | 2.16 (0.57,3.76) | Ref. | |
| 2 | 50.9 (41.5,60.2) | 1.08 (0.84,1.41) | 0.54 | 65.9 (49.0,82.9) | 1.58 (0.99,2.53) | 0.06 | 2.28 (0.99,3.58) | 1.06 (0.42,2.66) | 0.91 |
| 3 | 52.0 (46.7,57.2) | 1.11 (0.90,1.37) | 0.33 | 57.4 (46.1,68.6) | 1.37 (0.88,2.13) | 0.16 | 3.28 (2.21,4.36) | 1.52 (0.68,3.38) | 0.31 |
| 4 | 52.1 (49.0,55.2) | 1.11 (0.91,1.35) | 0.29 | 51.1 (45.0,57.2) | 1.23 (0.81,1.85) | 0.33 | 3.72 (2.97,4.47) | 1.72 (0.80,3.67) | 0.16 |
| 5 Most deprived | 57.7 (55.5,59.8) | 1.23 (1.02,1.49) | 0.03 | 64.7 (60.3,69.2) | 1.55 (1.04,2.31) | 0.03 | 3.97 (3.49,4.45) | 1.83 (0.87,3.85) | 0.11 |
| India | | | | | | | | | |
| 1 Least deprived | 47.7 (44.1,51.2) | Ref. | | 75.5 (66.1,84.9) | Ref. | | 0.77 (0.35,1.18) | Ref. | |
| 2 | 44.5 (41.6,47.3) | 0.93 (0.85,1.03) | 0.17 | 87.4 (78.6,96.2) | 1.16 (0.99,1.36) | 0.07 | 0.88 (0.51,1.24) | 1.14 (0.58,2.26) | 0.70 |
| 3 | 42.0 (39.7,44.2) | 0.88 (0.80,0.96) | 0.01 | 76.4 (70.2,82.6) | 1.01 (0.87,1.17) | 0.87 | 1.53 (1.14,1.92) | 1.99 (1.10,3.60) | 0.02 |
| 4 | 40.5 (38.7,42.3) | 0.85 (0.78,0.93) | 0.00 | 72.2 (67.5,77.0) | 0.96 (0.83,1.10) | 0.53 | 1.93 (1.58,2.27) | 2.51 (1.43,4.42) | <0.01 |
| 5 Most deprived | 50.4 (48.2,52.6) | 1.06 (0.97,1.15) | 0.20 | 83.7 (77.7,89.6) | 1.11 (0.96,1.28) | 0.15 | 2.06 (1.67,2.46) | 2.69 (1.53,4.76) | <0.01 |
| Nigeria | | | | | | | | | |
| 1 Least deprived | 42.7 (35.6,49.9) | Ref. | | 27.9 (16.8,39.0) | Ref. | | 1.00 (0.02,1.97) | Ref. | |
| 2 | 36.4 (31.0,41.7) | 0.85 (0.68,1.06) | 0.16 | 27.6 (17.4,37.8) | 0.99 (0.58,1.68) | 0.97 | 0.18 (-0.17,0.53) | 0.18 (0.02,1.62) | 0.13 |
| 3 | 34.9 (31.4,38.5) | 0.82 (0.67,0.99) | 0.04 | 30.3 (23.1,37.5) | 1.09 (0.68,1.72) | 0.72 | 0.57 (0.15,0.99) | 0.57 (0.17,1.93) | 0.37 |
| 4 | 36.4 (34.0,38.7) | 0.85 (0.71,1.02) | 0.08 | 28.6 (24.1,33.2) | 1.03 (0.67,1.57) | 0.91 | 0.49 (0.25,0.73) | 0.49 (0.16,1.46) | 0.20 |
| 5 Most deprived | 40.2 (38.2,42.1) | 0.94 (0.79,1.12) | 0.48 | 27.2 (23.8,30.6) | 0.97 (0.64,1.48) | 0.90 | 0.57 (0.37,0.77) | 0.57 (0.20,1.61) | 0.29 |
| Pakistan | | | | | | | | | |
| 1 Least deprived | 54.7 (48.8,60.7) | Ref. | | 62.7 (50.8,74.7) | Ref. | | 1.41 (0.53,2.29) | Ref. | |
| 2 | 58.4 (53.4,63.4) | 1.07 (0.93,1.22) | 0.36 | 58.4 (49.2,67.6) | 0.93 (0.73,1.19) | 0.57 | 3.20 (2.25,4.15) | 2.27 (1.14,4.50) | 0.02 |
| 3 | 58.8 (54.6,62.9) | 1.07 (0.94,1.22) | 0.28 | 62.2 (55.8,68.7) | 0.99 (0.80,1.23) | 0.94 | 3.10 (2.48,3.71) | 2.19 (1.15,4.20) | 0.02 |
| 4 | 66.0 (63.4,68.6) | 1.20 (1.07,1.35) | <0.01 | 57.7 (53.4,61.9) | 0.92 (0.75,1.13) | 0.41 | 3.33 (2.87,3.79) | 2.36 (1.25,4.44) | 0.01 |
| 5 Most deprived | 81.0 (78.9,83.0) | 1.48 (1.32,1.65) | <0.01 | 61.9 (58.7,65.0) | 0.99 (0.81,1.20) | 0.89 | 3.48 (3.14,3.82) | 2.46 (1.32,4.60) | <0.01 |
| Poland | | | | | | | | | |
| 1 Least deprived | 32.8 (30.2,35.3) | Ref. | | 77.4 (68.8,86.0) | Ref. | | 1.49 (0.98,2.01) | Ref. | |
| 2 | 34.6 (32.4,36.7) | 1.06 (0.96,1.17) | 0.29 | 70.4 (63.5,77.4) | 0.91 (0.79,1.05) | 0.21 | 1.83 (1.36,2.31) | 1.23 (0.80,1.87) | 0.34 |
| 3 | 33.7 (32.1,35.3) | 1.03 (0.94,1.13) | 0.54 | 72.9 (67.5,78.3) | 0.94 (0.83,1.07) | 0.38 | 2.63 (2.19,3.07) | 1.76 (1.21,2.56) | <0.01 |
| 4 | 34.1 (32.8,35.5) | 1.04 (0.96,1.14) | 0.36 | 75.7 (71.2,80.3) | 0.98 (0.86,1.11) | 0.74 | 2.72 (2.34,3.10) | 1.82 (1.26,2.61) | <0.01 |
| 5 Most deprived | 41.7 (40.2,43.2) | 1.27 (1.17,1.39) | <0.01 | 82.5 (77.9,87.1) | 1.07 (0.94,1.20) | 0.30 | 3.39 (2.96,3.81) | 2.27 (1.58,3.24) | <0.01 |
| UK | | | | | | | | | |
| 1 Least deprived | 55.7 (55.1,56.4) | Ref. | | 57.1 (55.8,58.4) | Ref. | | 0.54 (0.49,0.59) | Ref. | |

| | | | | | | | | | |
|---|---|---|---|---|---|---|---|---|---|
| 2 | 61.8 (61.2,62.5) | 1.00 (1.00,1.00) | <0.01 | 58.0 (56.7,59.3) | 1.02 (0.99,1.04) | 0.19 | 0.84 (0.78,0.91) | 1.56 (1.42,1.71) | <0.01 |
| 3 | 64.6 (63.9,65.3) | 0.00 (0.00,0.00) | <0.01 | 55.8 (54.6,57.1) | 0.98 (0.95,1.00) | 0.06 | 1.09 (1.02,1.17) | 2.02 (1.85,2.20) | <0.01 |
| 4 | 71.8 (71.1,72.5) | 0.00 (0.00,0.00) | <0.01 | 54.5 (53.3,55.7) | 0.95 (0.93,0.98) | <0.01 | 1.68 (1.57,1.79) | 3.10 (2.86,3.36) | <0.01 |
| 5 Most deprived | 84.2 (83.4,85.0) | 0.00 (0.00,0.00) | <0.01 | 51.6 (50.4,52.7) | 0.90 (0.88,0.92) | <0.01 | 2.82 (2.65,2.99) | 5.21 (4.82,5.62) | <0.01 |

CI = Confidence interval, IMD = index of multiple deprivation, IRR = incidence rate ratio; *results derived from negative binomial/Poisson regression models adjusted for year of birth, maternal region of birth, IMD group and maternal region of birth*IMD group interaction term (regression model results available on request); marginal incidence rates derived from models with year of birth set to mid-study (2011); IRR of admission rates for IMD groups in comparison to the least deprived IMD group, within maternal region groups; **Negative binomial regression, *N* = 3,492,139, AIC = 3685534.62 (compared with AIC = 3685728.14 for model without interaction term); ***Negative binomial regression, *N* = 3,492,139, AIC = 899210.58 (compared with AIC = 899254.24 for model without interaction term); [a]Poisson regression, *N* = 2,496,912, AIC = 168449.75 (compared with AIC = 168621.82 for model without interaction term)

**Supplementary Table 12. Estimated incidence rates (per 1000 child-years) and IRRs of hospital admissions with specified diagnoses, by migration status of parents and IMD group (derived from negative binomial/Poisson regression models)***

| | Acute infections** | | | Feeding difficulties and jaundice*** | | | Tooth extractions for caries[a] | | |
|---|---|---|---|---|---|---|---|---|---|
| | Incidence rate (95% CI) | IRR (95% CI) | p-value | Incidence rate (95% CI) | IRR (95% CI) | p-value | Incidence rate (95% CI) | IRR (95% CI) | p-value |
| Mother UK-born & SP UK-born | | | | | | | | | |
| 1 Least deprived | 55.7 (55.1, 56.3) | Ref. | | 57.4 (56.1, 58.7) | Ref. | | 0.54 (0.49, 0.59) | Ref. | |
| 2 | 61.6 (61.0, 62.3) | 1.11 (1.09, 1.12) | <0.01 | 58.5 (57.2, 59.8) | 1.02 (0.99, 1.05) | 0.13 | 0.82 (0.75, 0.88) | 1.52 (1.38, 1.68) | |
| 3 | 64.2 (63.5, 64.9) | 1.15 (1.14, 1.17) | <0.01 | 56.5 (55.2, 57.7) | 0.98 (0.96, 1.01) | 0.22 | 1.07 (0.99, 1.14) | 1.99 (1.81, 2.18) | <0.01 |
| 4 | 71.8 (71.0, 72.5) | 1.29 (1.27, 1.30) | <0.01 | 55.0 (53.8, 56.2) | 0.96 (0.94, 0.98) | <0.01 | 1.62 (1.52, 1.72) | 3.00 (2.75, 3.28) | <0.01 |
| 5 Most deprived | 83.8 (83.0, 84.6) | 1.50 (1.49, 1.52) | <0.01 | 51.7 (50.5, 52.8) | 0.90 (0.88, 0.92) | <0.01 | 2.81 (2.65, 2.96) | 5.22 (4.81, 5.68) | <0.01 |
| Mother UK-born & SP non-UK-born | | | | | | | | | |
| 1 Least deprived | 46.5 (44.7, 48.4) | Ref. | | 61.3 (57.0, 65.6) | Ref. | | 0.64 (0.46, 0.82) | Ref. | |
| 2 | 50.5 (48.6, 52.3) | 1.08 (1.03, 1.14) | <0.01 | 61.8 (57.6, 66.0) | 1.01 (0.92, 1.11) | 0.88 | 1.13 (0.89, 1.37) | 1.77 (1.25, 2.51) | <0.01 |
| 3 | 52.8 (51.0, 54.5) | 1.13 (1.08, 1.19) | <0.01 | 59.1 (55.3, 62.8) | 0.96 (0.88, 1.06) | 0.42 | 1.25 (1.02, 1.48) | 1.96 (1.41, 2.74) | <0.01 |
| 4 | 58.7 (56.9, 60.4) | 1.26 (1.20, 1.32) | <0.01 | 58.6 (55.4, 61.8) | 0.96 (0.88, 1.04) | 0.30 | 2.09 (1.82, 2.36) | 3.28 (2.42, 4.44) | <0.01 |
| 5 Most deprived | 76.5 (74.7, 78.2) | 1.64 (1.57, 1.72) | <0.01 | 60.9 (58.1, 63.7) | 0.99 (0.92, 1.08) | 0.88 | 3.15 (2.86, 3.45) | 4.95 (3.70, 6.62) | <0.01 |
| Mother UK-born (sole registration) | | | | | | | | | |
| 1 Least deprived | 78.3 (74.4, 82.2) | Ref. | | 59.1 (52.3, 65.9) | Ref. | | 1.68 (1.21, 2.14) | Ref. | |
| 2 | 85.0 (81.6, 88.4) | 1.09 (1.02, 1.16) | 0.01 | 54.8 (49.6, 60.0) | 0.93 (0.80, 1.07) | 0.31 | 2.35 (1.90, 2.80) | 1.40 (1.01, 1.95) | 0.04 |
| 3 | 85.7 (82.9, 88.5) | 1.09 (1.03, 1.16) | <0.01 | 50.0 (46.0, 54.1) | 0.85 (0.74, 0.97) | 0.02 | 2.47 (2.08, 2.85) | 1.47 (1.08, 2.01) | 0.02 |
| 4 | 85.2 (83.1, 87.3) | 1.09 (1.03, 1.15) | <0.01 | 50.8 (47.5, 54.0) | 0.86 (0.75, 0.98) | 0.02 | 3.09 (2.75, 3.44) | 1.84 (1.38, 2.47) | <0.01 |
| 5 Most deprived | 93.1 (91.3, 94.9) | 1.19 (1.13, 1.25) | <0.01 | 46.3 (44.0, 48.6) | 0.78 (0.69, 0.89) | <0.01 | 3.84 (3.52, 4.16) | 2.29 (1.72, 3.03) | <0.01 |
| Mother non-UK-born & SP non-UK-born | | | | | | | | | |
| 1 Least deprived | 36.9 (35.7, 38.1) | Ref. | | 74.4 (70.7, 78.1) | Ref. | | 1.26 (1.05, 1.47) | Ref. | |
| 2 | 35.8 (34.8, 36.8) | 0.97 (0.93, 1.01) | 0.19 | 70.8 (67.7, 73.9) | 0.95 (0.89, 1.01) | 0.13 | 1.84 (1.62, 2.07) | 1.46 (1.20, 1.78) | <0.01 |
| 3 | 37.5 (36.7, 38.4) | 1.02 (0.98, 1.06) | 0.37 | 67.2 (64.8, 69.6) | 0.90 (0.85, 0.96) | <0.01 | 2.49 (2.28, 2.71) | 1.98 (1.66, 2.36) | <0.01 |
| 4 | 40.0 (39.3, 40.7) | 1.09 (1.05, 1.12) | <0.01 | 63.2 (61.4, 65.1) | 0.85 (0.80, 0.90) | <0.01 | 3.04 (2.83, 3.25) | 2.41 (2.04, 2.85) | <0.01 |
| 5 Most deprived | 51.2 (50.5, 51.9) | 1.39 (1.34, 1.44) | <0.01 | 64.9 (63.2, 66.6) | 0.87 (0.83, 0.92) | <0.01 | 3.45 (3.24, 3.66) | 2.73 (2.32, 3.22) | <0.01 |
| Mother non-UK-born & SP UK-born | | | | | | | | | |
| 1 Least deprived | 41.3 (39.9, 42.6) | Ref. | | 62.5 (59.0, 66.0) | Ref. | | 0.73 (0.57, 0.89) | Ref. | |
| 2 | 43.1 (41.6, 44.5) | 1.04 (1.00, 1.09) | 0.07 | 65.9 (62.2, 69.6) | 1.05 (0.98, 1.14) | 0.18 | 0.96 (0.77, 1.15) | 1.31 (0.98, 1.76) | 0.07 |
| 3 | 44.4 (43.0, 45.9) | 1.08 (1.03, 1.13) | <0.01 | 64.4 (60.9, 67.9) | 1.03 (0.95, 1.11) | 0.45 | 1.24 (1.04, 1.45) | 1.70 (1.30, 2.23) | <0.01 |
| 4 | 48.2 (46.8, 49.6) | 1.17 (1.12, 1.22) | <0.01 | 63.7 (60.5, 66.9) | 1.02 (0.95, 1.10) | 0.62 | 1.93 (1.69, 2.18) | 2.65 (2.06, 3.40) | <0.01 |
| 5 Most deprived | 65.4 (63.7, 67.1) | 1.58 (1.52, 1.65) | <0.01 | 60.3 (57.3, 63.3) | 0.96 (0.90, 1.04) | 0.33 | 2.98 (2.68, 3.29) | 4.08 (3.22, 5.18) | <0.01 |
| Mother non-UK-born (sole registration) | | | | | | | | | |
| 1 Least deprived | 40.5 (33.0, 48.1) | Ref. | | 46.1 (26.9, 65.3) | Ref. | | 0.33 (-0.32, 0.98) | Ref. | |

| | | | | | | | | | |
|---|---|---|---|---|---|---|---|---|---|
| 2 | 44.6 (38.9, 50.4) | 1.10 (0.88, 1.38) | 0.40 | 48.3 (35.3, 61.4) | 1.05 (0.64, 1.72) | 0.85 | 3.43 (1.93, 4.92) | 10.31 (1.39, 76.62) | 0.02 |
| 3 | 44.9 (40.9, 48.9) | 1.11 (0.90, 1.36) | 0.33 | 54.1 (44.9, 63.4) | 1.17 (0.75, 1.84) | 0.48 | 2.90 (2.00, 3.80) | 8.73 (1.20, 63.36) | 0.03 |
| 4 | 44.0 (41.4, 46.6) | 1.09 (0.89, 1.32) | 0.40 | 42.1 (36.8, 47.4) | 0.91 (0.59, 1.41) | 0.68 | 3.86 (3.13, 4.60) | 11.63 (1.63, 83.10) | 0.01 |
| 5 Most deprived | 52.5 (50.1, 54.8) | 1.30 (1.07, 1.57) | 0.01 | 42.6 (38.4, 46.9) | 0.92 (0.60, 1.42) | 0.72 | 3.92 (3.35, 4.48) | 11.79 (1.66, 83.92) | 0.01 |

CI = Confidence interval, IMD = index of multiple deprivation, IRR = incidence rate ratio, SP=second parent; * results derived from negative binomial/Poisson regression models adjusted for year of birth, parental migration status, IMD group and parental migration status*IMD group interaction term (regression model results available on request); marginal incidence rates derived from models with year of birth set to mid-study (2011); IRR of admission rates for IMD groups in comparison to the least deprived IMD group, within parental migration status group; **negative binomial regression, *N* = 4,174,596, AIC = 4206508.56 (compared with AIC = 4206994.09 for model without interaction term); ***Negative binomial regression, *N* = 4,174,596, AIC = 4206508.56 with interaction term, AIC = 4206994.09without interaction term; [a]Poisson regression, *N* = 2,973,284, AIC = 220,264 with interaction term, AIC = 220,433 without interaction term

#### 7. Appendix G. Results: sensitivity analysis

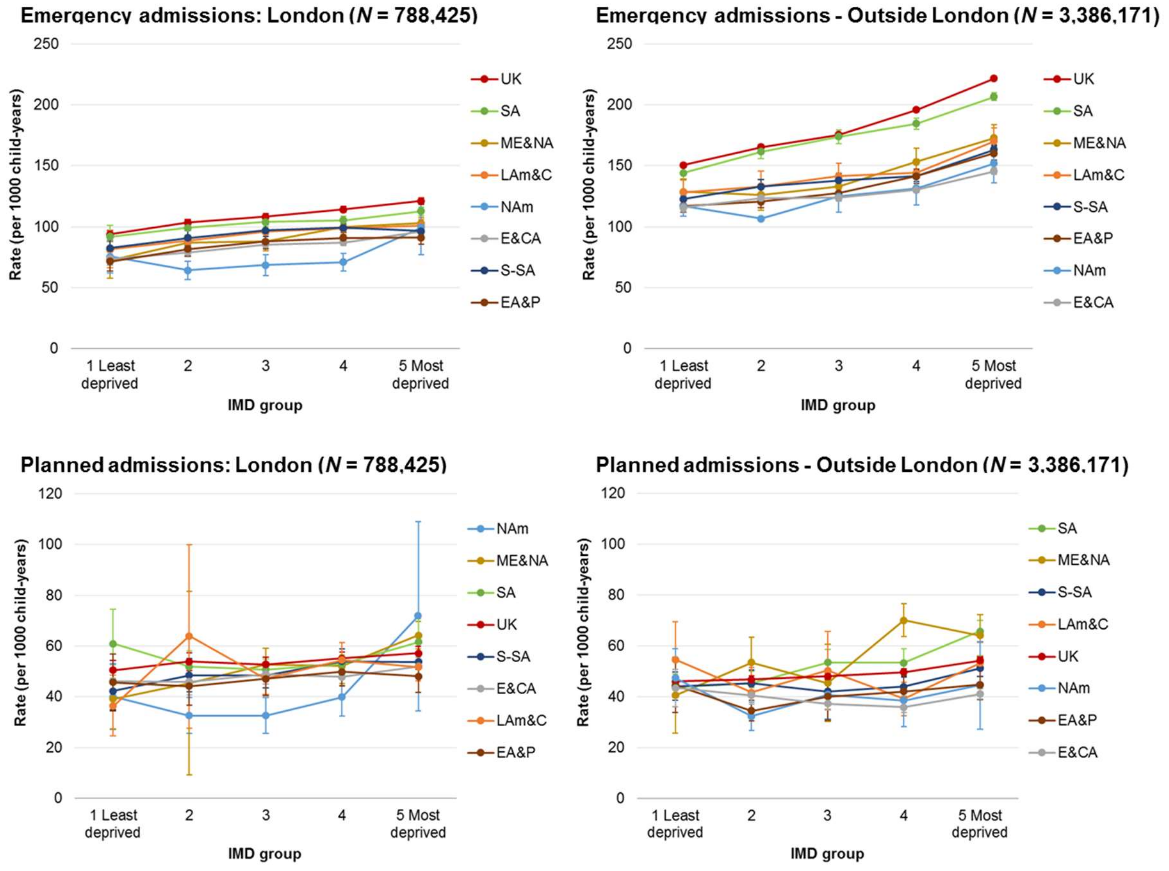


**Supplementary Figures 5. Estimated incidence rates* of emergency and planned hospital admissions (per 1000 person-years), by maternal region of birth and IMD group: stratified by London/non-London residence at birth.** N = number of children included in each analyses; IMD= index of multiple deprivation; EA&P= East Asia and Pacific, E&CA= Europe (excl. UK) and Central Asia, LAm&C= Latin America and Caribbean, ME&NA= Middle East and North Africa, NAm= North America, SA= South Asia, S-SA= Sub-Saharan Africa; *with year of birth set to mid-study (2011) derived from negative binomial regression models adjusted for year of birth, maternal world region of birth, IMD group, and maternal world region of birth*IMD group interaction term.

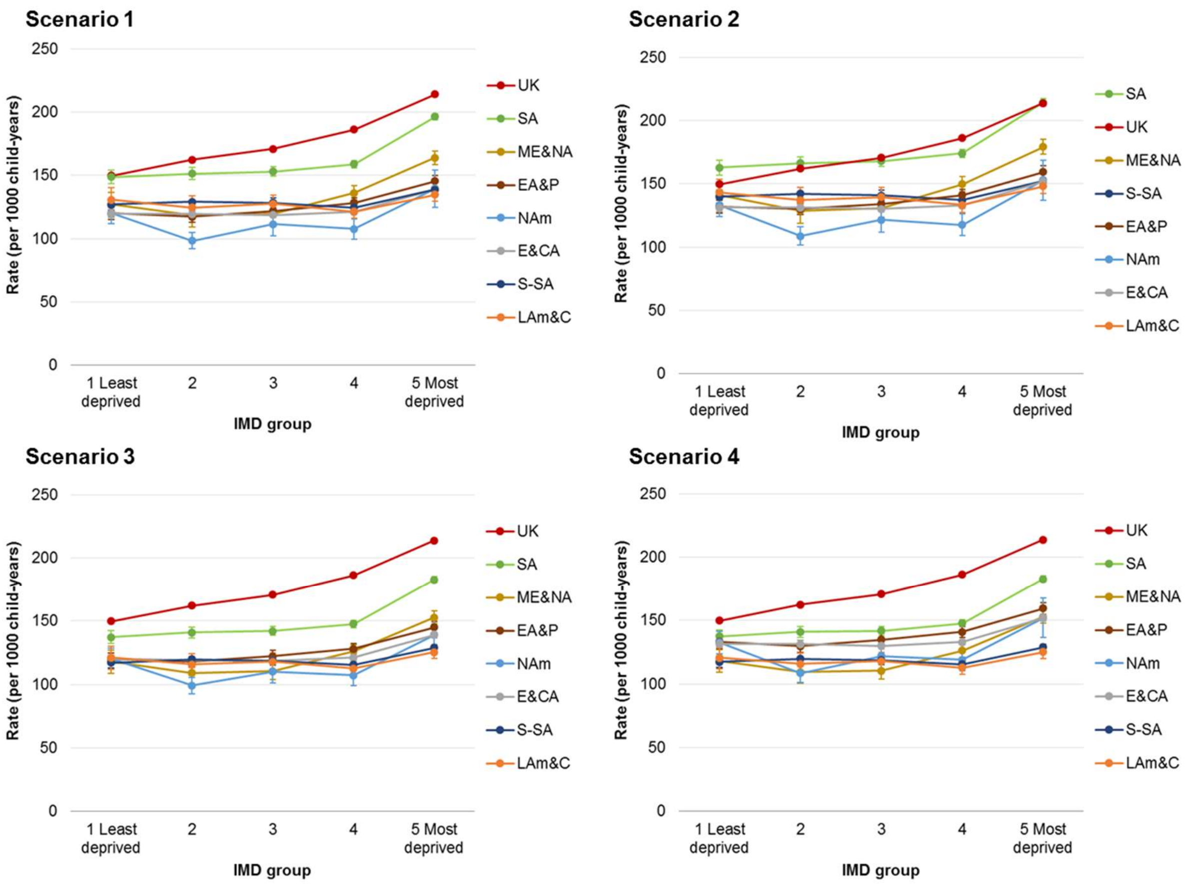


**Supplementary Figures 6. Estimated incidence rates* of emergency admissions to hospital (per 1000 child-years), by maternal region of birth: stratified by emigration sensitivity analysis scenario (see Appendix C).** IMD= index of multiple deprivation; EA&P= East Asia and Pacific, E&CA= Europe (excl. UK) and Central Asia, LAm&C= Latin America and Caribbean, ME&NA= Middle East and North Africa, NAm= North America, SA= South Asia, S-SA= Sub-Saharan Africa; *with year of birth set to mid-study (2011) derived from negative binomial regression models (*N* = 4,174,596) adjusted for year of birth, maternal world region of birth, IMD group, and maternal world region of birth*IMD group interaction term.

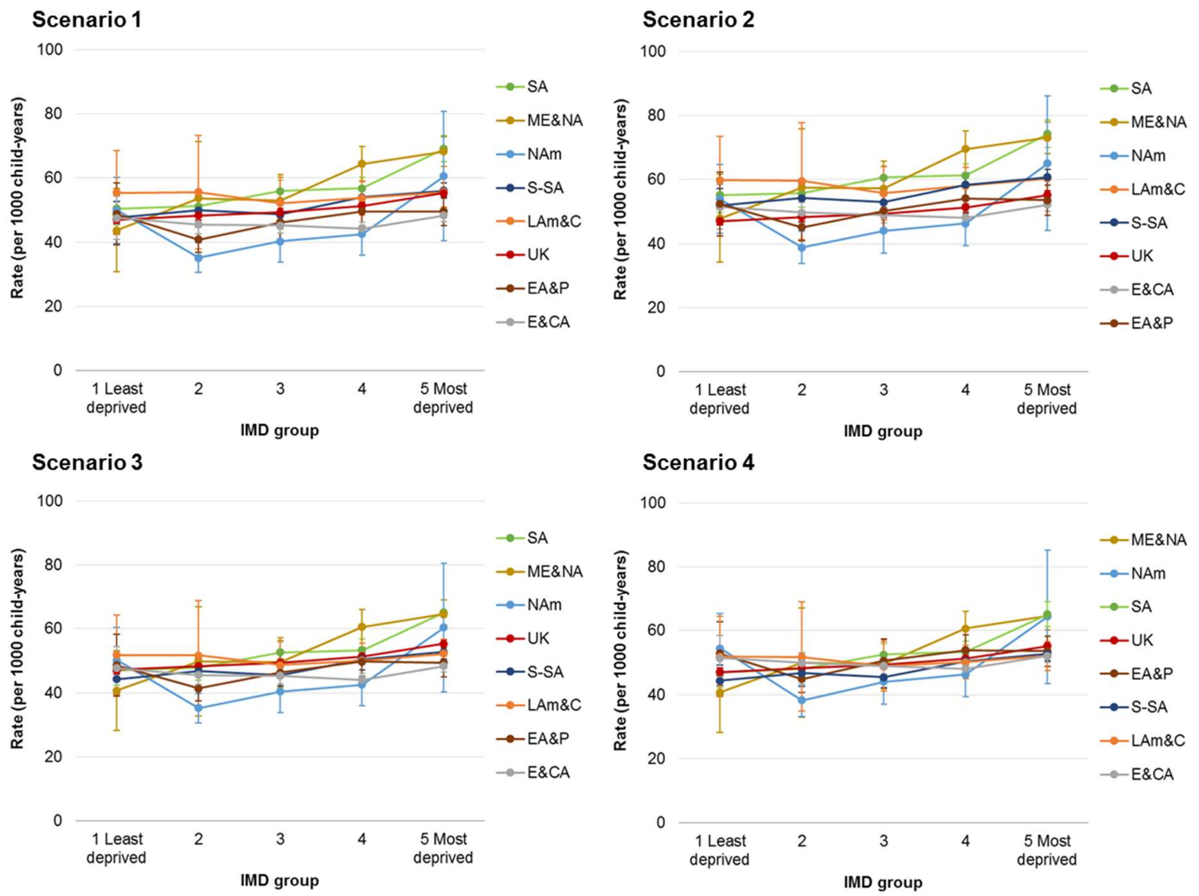


**Supplementary Figures 7. Estimated incidence rates* of planned admissions to hospital (per 1000 child-years), by maternal region of birth: stratified by emigration sensitivity analysis scenario (see Appendix C).** IMD= index of multiple deprivation; EA&P= East Asia and Pacific, E&CA= Europe (excl. UK) and Central Asia, LAm&C= Latin America and Caribbean, ME&NA= Middle East and North Africa, NAm= North America, SA= South Asia, S-SA= Sub-Saharan Africa; *with year of birth set to mid-study (2011) derived from negative binomial regression models ($N$ = 4,174,596) adjusted for year of birth, maternal world region of birth, IMD group, and maternal world region of birth*IMD group interaction term.

#### 8. Supplementary material references

1 Gill PJ, Goldacre MJ, Mant D, *et al.* Increase in emergency admissions to hospital for children aged under 15 in England, 1999–2010: national database analysis. *Arch Dis Child* 2013; **98**: 328.

2 NHS Digital. NHS Outcomes Framework Indicators - February 2021 Release. 2021. https://digital.nhs.uk/data-and-information/publications/statistical/nhs-outcomes-framework/february-2021#chapter-index.

3 Flaherman V, Schaefer EW, Kuzniewicz MW, Li SX, Walsh EM, Paul IM. Health Care Utilization in the First Month After Birth and Its Relationship to Newborn Weight Loss and Method of Feeding. *Academic Pediatrics* 2018; **18**: 677–84.

4 Zylbersztejn A, Verfürden M, Hardelid P, Gilbert R, Wijlaars L. Phenotyping congenital anomalies in administrative hospital records. *Paediatric and Perinatal Epidemiology* 2020; **34**: 21–8.

5 Schisterman EF, Cole SR, Platt RW. Overadjustment bias and unnecessary adjustment in epidemiologic studies. *Epidemiology* 2009; **20**: 488–95.

6 Katikireddi SV, Cezard G, Bhopal RS, *et al.* Assessment of health care, hospital admissions, and mortality by ethnicity: population-based cohort study of health-system performance in Scotland. *The Lancet Public Health* 2018; **3**: e226–36.

7 Office for National Statistics. Annual population survey (APS) QMI. 2012. https://www.ons.gov.uk/employmentandlabourmarket/peopleinwork/employmentandemployeetypes/methodologies/annualpopulationsurveyapsqmi.

8 Office for National Statistics. Long-Term International Migration estimates methodology. 2020. https://www.ons.gov.uk/peoplepopulationandcommunity/populationandmigration/internationalmigration/methodologies/longterminternationalmigrationestimatesmethodology.

9 Office for National Statistics. International Passenger Survey: quality information in relation to migration flows. 2018. https://www.ons.gov.uk/peoplepopulationandcommunity/populationandmigration/internationalmigration/methodologies/internationalpassengersurveyqualityinformationinrelationtomigrationflows.

10 Office for National Statistics. Note on the differences between Long-Term International Migration flows derived from the International Passenger Survey and estimates of the population obtained from the Annual Population Survey. 2016. https://www.ons.gov.uk/peoplepopulationandcommunity/populationandmigration/internationalmigration/articles/noteonthedifferencesbetweenlongterminternationalmigrationflowsderivedfromtheinternationalpassengersurveyandestimatesofthepopulationobtainedfromtheannualpopulationsurvey/december2016.

11 Home Office. Research and analysis: Emigration from the UK. 2012. https://www.gov.uk/government/publications/emigration-from-the-uk.
